# Monitoring ESBL-*Escherichia coli* in Swiss wastewater between November 2021 and November 2022: insights into population carriage

**DOI:** 10.1101/2023.11.12.23298428

**Authors:** Sheena Conforti, Aurélie Holschneider, Émile Sylvestre, Timothy R. Julian

**Affiliations:** Eawag, Swiss Federal Institute of Aquatic Science and Technology, Dübendorf CH-8600, Switzerland; Department of Biosystems Science and Engineering, ETH Zürich, 4051 Basel, Switzerland; Swiss Tropical and Public Health Institute, 4051 Basel, Switzerland; University of Basel, 4055 Basel, Switzerland

## Abstract

**Background:** Antimicrobial resistance (AMR) poses a global health threat, causing millions of deaths annually, with expectations of increased impact in the future. Wastewater surveillance offers a cost-effective, non-invasive tool to understand AMR carriage trends within a population.

**Aim:** We monitored extended-spectrum β-lactamase producing *Escherichia coli* (ESBL-*E. coli*) weekly in influent wastewater from six wastewater treatment plants (WWTPs) in Switzerland (November 2021 to November 2022) to investigate spatio-temporal variations, explore correlations with environmental variables, develop a predictive model for ESBL-*E. coli* carriage in the community, and detect the most prevalent ESBL-genes.

**Methods:** We cultured total and ESBL-*E. coli* in 300 wastewater samples to quantify daily loads and percentage of ESBL-*E. coli*. Additionally, we screened 234 ESBL-*E. coli* isolates using molecular-methods for the presence of 18 ESBL-gene families.

**Results:** We found a population-weighted mean percentage of ESBL-*E. coli* of 1.9% (95%CI 1.8%, 2%) across all sites and weeks, which can inform ESBL-*E. coli* carriage. Concentrations of ESBL-*E. coli* varied across WWTPs and time, with higher values observed in WWTPs serving larger populations. Recent precipitations (previous 24-/96-hours) showed no significant association with ESBL-*E. coli*, while temperature occasionally had a moderate impact (p<0.05, correlation coefficients approximately 0.40) in some locations. We identified *bla*_CTX-M-1_, *bla*_CTX-M-9_ and *bla*_TEM_ as the predominant ESBL-gene families.

**Conclusions:** Our study demonstrates that wastewater-based surveillance of culturable ESBL-*E. coli* provides insights into AMR trends in Switzerland and may also inform resistance. These findings establish a foundation for long-term, nationally established monitoring protocols and provide information that may help inform targeted public health interventions.

## Introduction

Antimicrobial resistance (AMR) is listed as one of the top ten most serious global health threats by the World Health Organization [1]. In 2019, an estimated five million deaths globally were associated with AMR infections [2]. Among the factors contributing to these fatalities is the transmission of AMR, with environmental pathways playing a significant role in both the spread of antibiotic-resistant bacteria (ARB) and the acquisition of antibiotic-resistant genes (ARGs) by clinically relevant pathogens [3].

ESBL-*E. coli* are resistant to clinically important antimicrobial drugs and are considered important vectors in the transmission of ARGs [4]. ESBL-*E. coli* are transmissible through human and animal faecal pollution and are commonly found in inflow wastewater worldwide [5-8]. Wastewater monitoring provides a comprehensive perspective of AMR carried across a human population [9,10], and it is more advantageous than the current patient sampling since it avoids privacy and ethical concerns related to the collection of patient data [11]. A centralized monitoring of AMR in wastewater can reflect community-level dissemination of AMR, providing understanding of the evolution and the spread of clinically relevant ARBs [11]. A previous study on carbapenemase-producing *E. coli* (CPE) conducted in Dutch municipal wastewater showed its sensitivity in estimating CPE prevalence in humans [12], suggesting that wastewater monitoring can inform prevalence rates and dynamics of other fecally shed antimicrobial resistant bacteria such as ESBL-*E. coli*.

Recently, ESBL-producing *Enterobacteriaceae* were monitored on a monthly scale in municipal wastewater of Basel, Switzerland [13]. Building on this, a centralized wastewater monitoring system of AMR and ARBs in the Swiss population may enable public health officials to better understand challenges associated with antimicrobial resistance and to target interventions more effectively. Previous studies showed that antibiotic resistance prevalence in bacteria from wastewater follows similar trends to the rates monitored in clinical environments [5,6,8,14].

Trends in ESBL-*E. coli* prevalence in wastewater are likely reflective of trends in ESBL-*E. coli* carriage in the population served. That is, concentrations of ESBL-*E. coli* in wastewater are a function of the prevalence of people carrying ESBL-*E. coli* in the catchment, as well as the proportion of ESBL-*E. coli* among all *E. coli* shed by carriers. However, concentrations may also be influenced by in sewer fate and transport processes, including temperature dependent decay and potential non-human sources of ESBL-*E. coli* entering combined sewers from urban water run-off [15]. Temporal trends in ESBL-*E. coli* concentrations may therefore reflect changes in carriage within the population, changes in shedding load dynamics, or environmental impacts on fate and transport. Previous studies of ESBL-*E. coli* prevalence report both, significant variations in ESBL-*E. coli* prevalence between months [13,16] as well as relatively stable percentages over time [6,4,17,18].

In this one-year longitudinal study, we monitored ESBL-*E. coli* in Swiss municipal wastewater weekly from six WWTPs. Using culture-based methods, we investigated national spatio-temporal variations of ESBL-*E. coli*, developed a predictive model for ESBL-*E. coli* carriage, explored ESBL-*E. coli* correlations with environmental variables, and assessed monitoring frequency impact on ESBL-*E. coli* concentrations estimation. Molecular methods characterized the presence of ESBL genes in 234 isolates. Our findings offer insights into ESBL-*E. coli* prevalence, distribution, and associated genes in Swiss wastewater, and may act as a basis for long-term wastewater-based surveillance of AMR.

## Methods

### Samples collection

Wastewater samples were collected from six wastewater treatment plants (WWTPs) in Switzerland serving an estimated 1.23 million residents (14% of the population): ARA Altenrhein (64,000 residents), ARA Chur (55,000 residents), STEP d’Aïre Genève (454,000 residents), ARA Sensetal Laupen (62,000 residents), IDA CDA Lugano (124,000 residents), and ARA Werdhölzli Zürich (471,000 residents) (Figure 1). Samples were collected weekly over a 1-year period from November 16, 2021, to November 29, 2022 (Supplementary Table S1). Wastewater was sampled using 24-hr flow-proportional composite sampling and transported to Eawag (Dübendorf, CH) in high-density polyethylene bottles with ice packs, stored at 4°C and processed within 48 hours.

**Figure 1:**
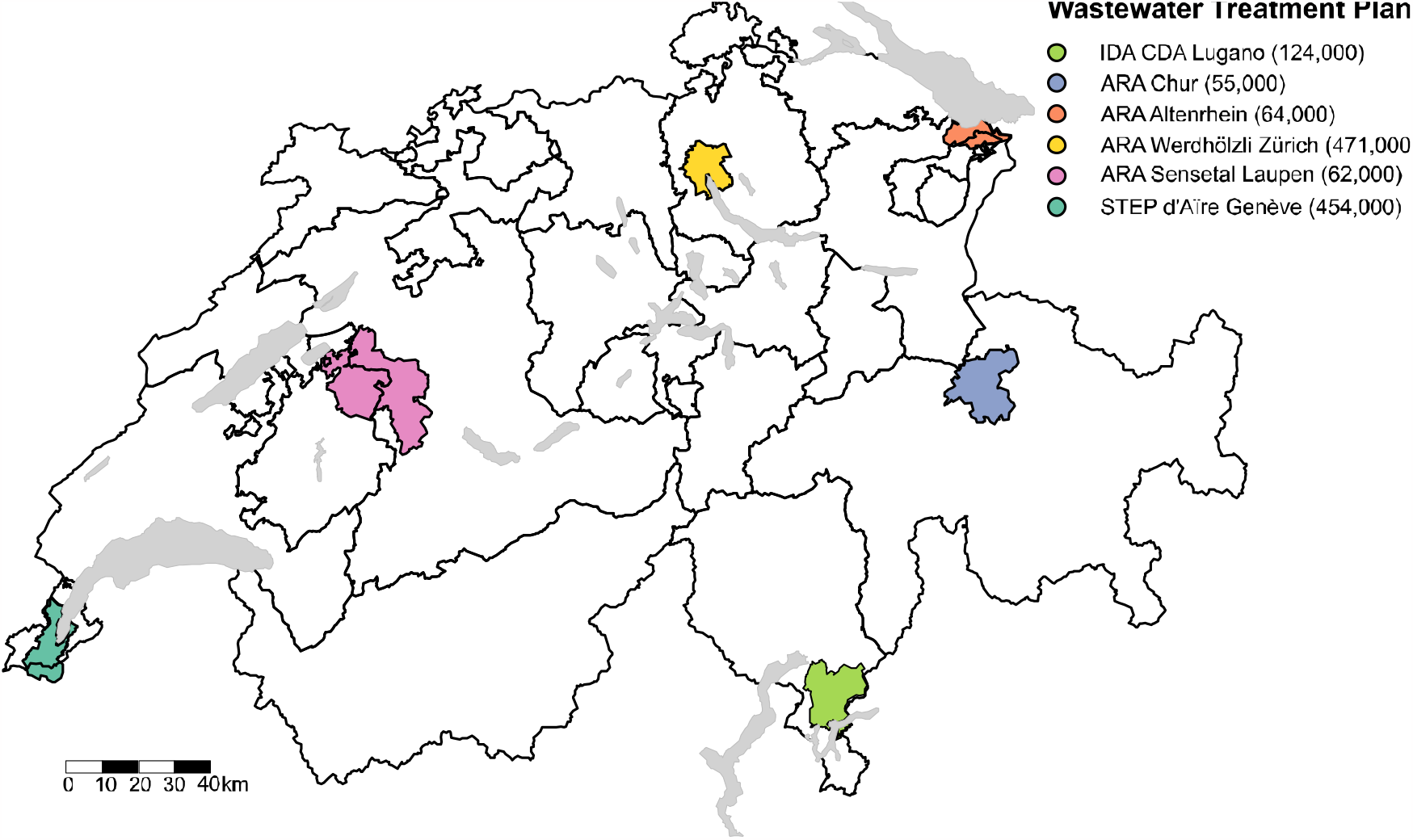
Geographic distribution, catchment area, and number of people served at each of the six investigated wastewater treatment plants in Switzerland.

### Enumeration of total and ESBL-*E. coli*

To enumerate total *E. coli*, wastewater samples were serially diluted 100-fold with sterile 0.9% NaCl and 100μl were plated on CHROMagar Orientation chromogenic media (CHROMagar™, bio-Mérieux, Marcy-l’Étoile, France). To enumerate ESBL-*E. coli*, 100μl of undiluted wastewater samples were plated on CHROMagar ESBL chromogenic media (CHROMagar™ ESBL, bio-Mérieux, Marcy-l’Étoile, France). Samples were plated in single replicates until February 08, 2022, and afterward in duplicates. Plates were incubated at 37°C for 24 hours. Then, colony concentrations were determined by counting the dark pink to reddish colonies on the CHROMagar Orientation for total *E. coli* and CHROMagar ESBL for ESBL-*E. coli*.

### Isolation, culturing and DNA extraction of ESBL-E. coli

A subset of presumptive ESBL-*E. coli* colonies were isolated every four weeks from CHROMagar ESBL plates. Specifically, three colonies per WWTP were streaked on LB Agar (Lennox) and incubated at 37°C for 20-24 hours. Single colonies were liquid cultured in Luria Broth (AppliChem), and DNA was extracted from 100μl of enriched liquid culture by boiling it at 100°C for one hour. DNA was stored at -20°C for subsequent analyses.

### Screening of ESBL-genes in ESBL-*E. coli* using dMLA

A total of 234 ESBL-*E. coli* isolates underwent screening for 18 ESBL-gene families using the digital Multiplex Ligation Assay (dMLA) [19]. Three screenings were conducted to cover all isolates, each including three no template controls (NTC) with DNA-/RNAse free water. The 72 half-probes used in the dMLA were mixed to achieve a final concentration of 1μM each. Subsequent ligation and PCR followed the previously described method [19] and were carried out in 96-well PCR Skirted plates (Milian) sealed with SilverSeal Sealer (Greiner Bio-One GmbH). Each well contained only one isolate or control sample. The PCR products (4μL from each), barcoded during the dMLA protocol, were pooled and purified using the Wizard® Genomic DNA Purification Kit. The pooled purified PCR products were then sent for Illumina MiSeq sequencing (Eurofins Genomics, GmbH, Ebersberg, Germany) with 150bp paired-end reads.

### dMLA sequence data analysis

NGS reads in FASTQ format were processed using Nextflow (Seqera Labs, v21.10.0.5640). The nsearch software [20] filtered reads with a minimum identity threshold of 0.8 and allowed one expected error per base pair. A Python (v3.9.2) code expanded degenerated sequences, retained matches to target probe-pairs, and classified reads by forward primer barcode and target probe-pair. Target quantification was performed in R (v4.1.1). To determine the detection limits for dMLA, false positive signals from the three NTC samples in each screening were analysed as previously reported [19]. To limit false positives, genes were only considered present if detectable by both probe-pairs, genes positive to one probe-pair were considered negative.

### Statistical analyses

All analyses were performed in R (v4.1.1) and R Studio (v2022.12.0.353). The loads (CFUs/(person-day)) of total- and ESBL-*E. coli* were calculated by normalizing estimated concentrations by the corresponding daily flow rate (m^3^/day) of the WWTP and the number of residents connected to the WWTP. For both the percentage of ESBL-*E. coli* and the loads, the average was computed when sample duplicates were available to both display and analyse the data. Statistical analysis used Kruskal-Wallis non-parametric tests with Dunn’s pairwise comparisons and Bonferroni adjustment to assess differences in ESBL-*E. coli* percentages and loads among WWTPs and across months within each WWTP.

The Spearman’s correlation coefficient was employed to assess the relationship between ESBL-*E. coli* percentage, total- and ESBL-*E. coli* loads, and the corresponding environmental variables (precipitation, temperature). An alpha value of 0.05 was used to determine significance of associations, with p-values corrected using the Bonferroni adjustment. Precipitation data (daily total and cumulative sum over 96 hours) and air temperature data (daily average from 6-18 UTC) were obtained from the Federal Office of Meteorology and Climatology MeteoSwiss (IDAWEB 1.3.5.0 ©2016MeteoSwiss).

The impact of monitoring frequency on estimating the mean ESBL*-E. coli* percentage was assessed using a Bayesian inference method [21]. Candidate models, gamma and log-normal distributions, were selected to describe temporal variations in ESBL-*E. coli* percentage, and their fit was compared using the Bayesian Deviance Information Criterion. In comparing candidate models, the gamma distribution generally provided a better fit than the log-normal distribution at most sites, leading to its selection for evaluating percentages (Supplementary Table S2). The standard error of the mean (SE) of the gamma distribution was calculated as 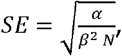, where *α* represents the shape parameter, *β* the rate parameter, and *N* the sample size. Subsequently, the 95% confidence interval (CI) of the mean, indicating the uncertainty of estimates, was calculated using the z-distribution.

ESBL-*E. coli* percentage in wastewater is conceptually related to the prevalence of ESBL-*E. coli* carriers within the domestic population contributing to the wastewater. Prevalence of ESBL-*E. coli* carriers is estimated by the ESBL-*E. coli* percentage in wastewater, denoted as *π*_*WW*_, divided by the average proportion of ESBL-*E. coli* relative to the total *E. coli* shed only by carriers in the population, denoted as 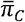, (Equation 1). In this study, 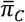 is not quantified, so the range of prevalence rates is modelled as a function of 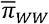.

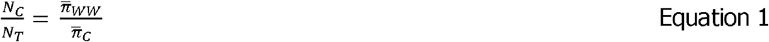

This model assumes constant 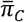, N_C_, and N_T_ over the data collection period for estimating 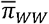. The total population (N_T_) includes only people colonized with *E. coli*, so is less than or equal to the total catchment population, considering those who do not shed *E. coli*. The model neglects fate and transport processes within the sewer system and during sample storage and transport. 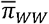 and 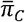 are estimated using the arithmetic mean. The model was applied for each WWTP, where 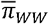 and its 95% CI were predicted from the gamma distribution (refer to Section “Impact of monitoring frequency on mean ESBL-*E. coli* percentage estimate”) and for combined WWTPs, using a population-weighted mean. The 95% CI was computed using the SE of the weighted mean, calculated using the method proposed by Endlich et *al*. [22], and the z-distribution.

## Results

Total *E. coli* were detectable in all samples with median load value of 1.4x10^10^ (IQR=1.4x10^10^). Similarly, ESBL-*E. coli* were always detectable with median 2.1x10^8^ (IQR=1.9x10^8^) CFUs/(person-day). The ESBL-*E. coli* median percentage was 1.6% (IQR=0.89%), and the population-adjusted averaged ESBL-*E. coli* percentage was 1.9% (±95% CI 0.1%). More information on summary statistics can be seen in Supplementary Table S3. ESBL-*E. coli* percentages significantly differed among WWTPs (p < 0.001), with the highest median percentage in Geneva (2.1% ;IQR=1.0%) and the lowest in Sensetal-Laupen (1.3;IQR=0.7%) (Figure 2). No significant monthly variations in ESBL-*E. coli* percentages were detected within any WWTPs (Supplementary Figure S1, Supplementary Tables S4, S5). Total *E. coli* loads differed significantly among WWTPs (p < 0.001), with the highest median loads in Altenrhein (2.3x10^10^ CFUs/person-day;IQR=1.4x10^10^), and the lowest in Lugano (8.8x10^9^ CFUs/person-day;IQR=4.6x10^9^) (Figure 2). Monthly variations were observed in Zurich (p < 0.001), Sensetal-Laupen (p < 0.05), Geneva (p < 0.001), and Altenrhein (p < 0.05). ESBL-*E. coli* loads varied significantly among WWTPs (p < 0.001), with the highest median loads in Altenrhein (3.3x10^8^ CFUs/person-day;IQR=2.3x10^8^), and the lowest in Lugano (1.7x10^8^ CFUs/person-day;IQR=1.6x10^8^), mirroring the pattern observed for total *E. coli* loads (Figure 2). Monthly variations were observed within Zurich (p < 0.001), Sensetal-Laupen (p < 0.001), and Geneva (p < 0.001). Notably, STEP d’Aïre Genève had significantly higher ESBL-*E. coli* loads in November 2022 compared to November 2021 (p < 0.05) and March 2022 (p < 0.05).

**Figure 2:**
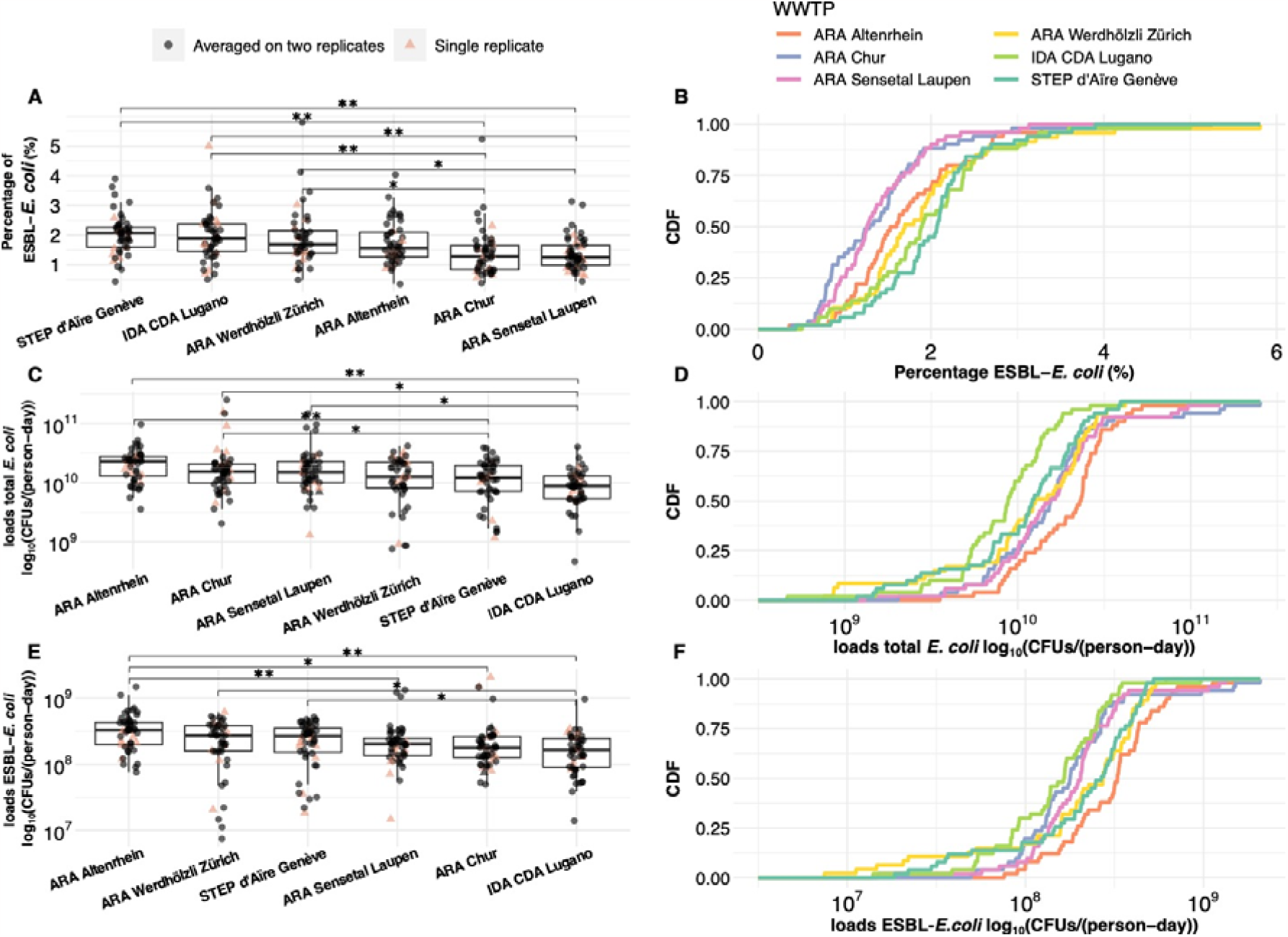
Comparative analysis of percentage and loads of extended-spectrum β-lactamase-producing *Escherichia coli* isolates among six wastewater treatment plants in Switzerland from November 2021 – November 2022. CDF: cumulative distribution function; CFUs: colony forming units; *E. coli: Escherichia coli*; ESBL: extended-spectrum β-lactamase; WWTP: wastewater treatment plant; (A) the percentage of ESBL-*E. coli* over total *E. coli* by WWTP, (B) the CDF of the percentage of ESBL-*E. coli* over total *E. coli* in each WWTP. (C) the log_10_ transformed loads of total *E. coli* by WWTP, and (D) the CDF of the log_10_ transformed loads of total *E. coli* in each WWTP. (E) the log_10_ transformed loads of ESBL-*E. coli* by WWTP, and (F) shows the CDF of the log_10_ transformed ESBL-*E. coli* loads in each WWTP. In (A), (C) and (E) black circles represent the average of duplicate values, while salmon triangles represent a single value where duplicates were unavailable. * indicates a significant difference with p<0.02 and ** with p<0.001.

We established a mechanistic relationship between estimated ESBL-*E. coli* percentage in wastewater and the average percentage of ESBL-*E. coli* in carriers’ guts (Figure 3). Considering the observed 1.9% population-averaged ESBL-*E. coli* percentage in Swiss wastewater, our model indicates prevalence between 1.9% (assuming all shed *E. coli* is ESBL-*E. coli*) and 100% (assuming 1.9% of all shed *E. coli* is ESBL-*E. coli*) (Table 1). At the European average ESBL-*E. coli* prevalence of 6% [23], approximately 32% of *E. coli* shed by carriers in Switzerland are ESBL-*E. coli*. Notably, if we consider the average ESBL-*E. coli* proportion in the gut of children in Bangladesh (19.3%) [24], ESBL-*E. coli* carriage prevalence in Switzerland varies between 9.2% and 10.5%. Assuming the consistent ESBL-*E. coli* proportion in carriers, observed differences in ESBL-*E. coli* percentages among WWTPs imply varying carriage rates. Estimated carriage rates range from 7.1% (95% CI 6.3%, 7.9%) in Sensetal-Laupen to 10.5% (95% CI 9.5%, 11.4%) in Geneva (Supplementary Figures S2 and S3).

**Figure 3:**
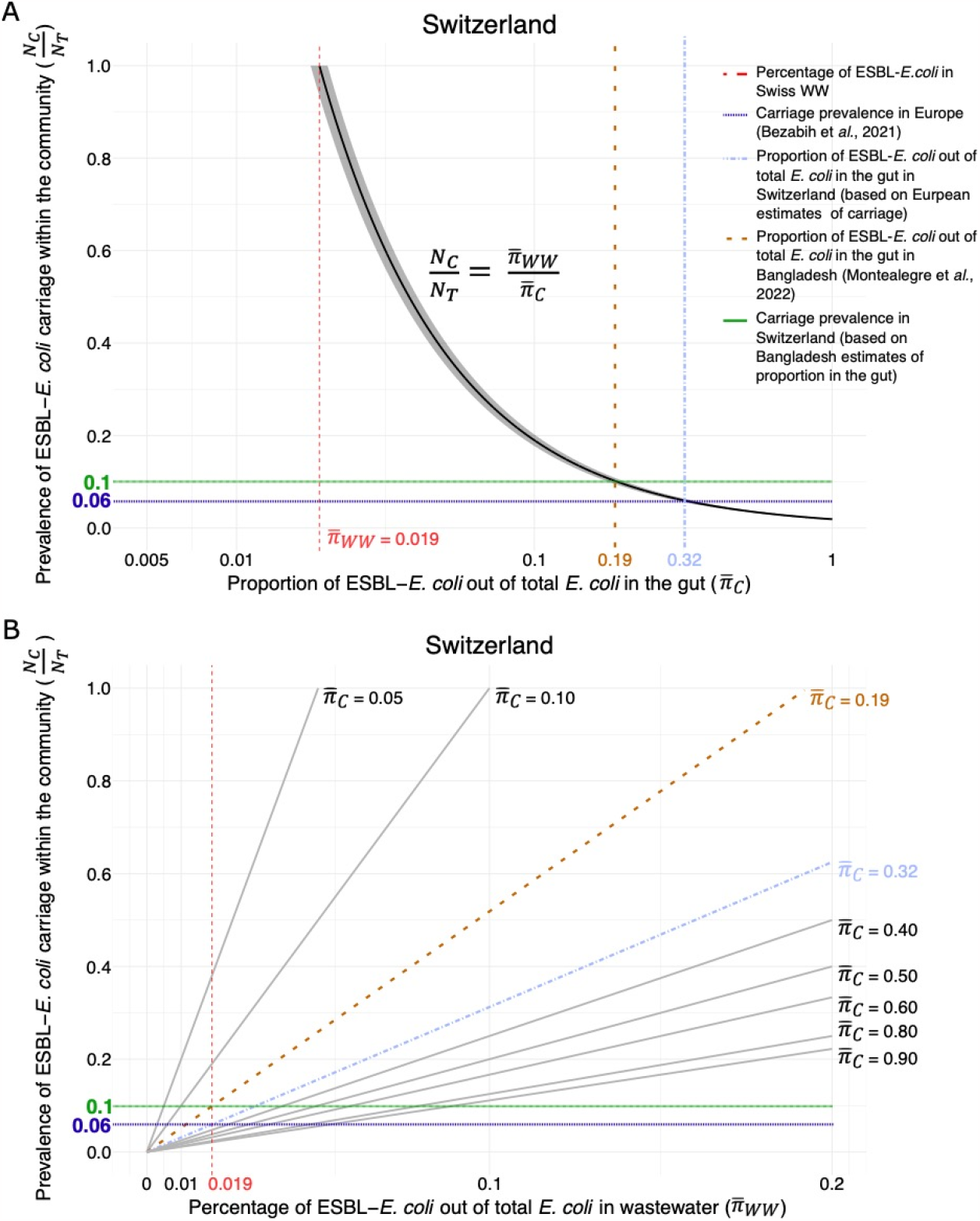
Relationship between percentage of extended-spectrum β-lactamase-producing *Escherichia coli* isolates in wastewater, prevalence of carriage within the community and proportion in the gut in Switzerland. *E. coli: Escherichia coli*; ESBL: extended-spectrum β-lactamase; *N*_*C*_ : prevalence of ESBL-*E. coli* carriers; *N*_*T*_ : total population size; 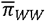: mean percentage of ESBL-*E. coli* in wastewater; 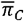: mean proportion of ESBL-*E. coli* relative to total *E. coli* in the gut discharged by carriers into wastewater (A) visualization of the relationship between percentage of ESBL-*E. coli* in wastewater 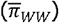.and prevalence of carriage within the community based on estimates of ESBL-*E. coli* out of total *E. coli* in the gut (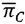, x-axis) equal to the percentage of ESBL-*E. coli* in wastewater. If the proportion of ESBL-*E. coli* in the gut 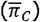 equals the percentage of ESBL-*E. coli* in wastewater 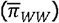, then 100% of the community shedding *E. coli* carries ESBL-*E. coli* (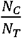, y-axis). The grey-shaded area on the graph represents the 95% confidence interval of the population-weighted mean percentage of ESBL-*E. coli* in wastewater. (B) describes the same relationship based on the percentage of ESBL-*E. coli* in wastewater 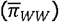 as a predictor (x-axis). Here, multiple values of 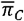 are assumed (straight lines), and the prevalence of ESBL-*E. coli* carriage within the community (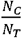, y-axis) can be inferred based on the percentage of ESBL-*E. coli* in wastewater 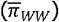. The dark-blue dotted line (6%) reflects an estimation by Bezabih et al. (2021) [23] for ESBL-*E. coli* carriage within Europe. Assuming this estimation holds for Switzerland, it corresponds to a 32% ESBL-*E. coli* proportion in the gut (light-blue dotted line). Alternatively, if the ESBL-*E. coli* proportion in the gut is approximately 19% (brown dotted line), as estimated in children from Bangladesh [31], then the carriage prevalence in Switzerland is approximately 10% (green line).

**Table 1:** Relationship between prevalence of ESBL-*E. coli* carriers and ESBL-*E. coli* percentage in wastewater. The table displays the estimated ESBL-*E. coli* carriage prevalence in the community as a function of the ESBL-*E. coli* proportion in the gut and the ESBL-*E. coli* percentage in wastewater. To describe the relationship within each wastewater treatment plant, the gamma arithmetic mean of the wastewater percentage was used. To describe the relationship across all WWTPs (Switzerland), the population-weighted mean was used.

| Average proportion of ESBL-*E. coli* out of total *E. coli* in the gut (%) | Prevalence of ESBL-*E. coli* carriage within the community (%) (95% CI of the mean) |  |  |  |  |  |  |
| --- | --- | --- | --- | --- | --- | --- | --- |
|  | Switzerland | ARA Werdhölzli Zürich | STEP d'Aire Genève | IDA CDA Lugano | ARA Altenrhein | ARA Sensetal Laupen | ARA Chur |
| 1.9 | 100<br>(93.8 - 100) | 100<br>(87.4 - 100) | 100<br>(96.8 - 100) | 100<br>(91.8 - 100) | 90.7<br>(80.4 - 100) | 72.3<br>(64.3 - 80.3) | 73.5<br>(62.1 - 85) |
| 10 | 18.9<br>(17.7 - 20.2) | 19.1<br>(16.6 - 21.6) | 20.2<br>(18.4 - 22) | 19.7<br>(17.4 - 22) | 17.2<br>(15.3 - 19.2) | 13.7<br>(12.2 - 15.2) | 14<br>(11.8 - 16.1) |
| 19.3<sup>a</sup> | 9.9<br>(9.2 - 10.5) | 9.9<br>(8.6 - 11.2) | 10.5<br>(9.5 - 11.4) | 10.2<br>(9 - 11.4) | 8.9<br>(7.9 - 10) | 7.1<br>(6.3 - 7.9) | 7.3<br>(6.1 - 8.4) |
| 30 | 6.3<br>(5.9 - 6.7) | 6.4<br>(5.5 - 7.2) | 6.7<br>(6.1 - 7.3) | 6.6<br>(5.8 - 7.3) | 5.7<br>(5.1 - 6.4) | 4.6<br>(4.1 - 5.1) | 4.7<br>(3.9 - 5.4) |
| 40 | 4.8<br>(4.4 - 5.1) | 4.8<br>(4.2 - 5.4) | 5.1<br>(4.6 - 5.5) | 4.9<br>(4.4 - 5.5) | 4.3<br>(3.3 - 4.8) | 3.4<br>(3.1 - 3.8) | 3.5<br>(3 - 4) |
| 50 | 3.8<br>(3.6 - 4.1) | 3.8<br>(3.3 - 4.3) | 4<br>(3.7 - 4.4) | 3.9<br>(3.56 - 4.4) | 3.4<br>(3.1 - 3.8) | 2.8<br>(2.4 - 3.1) | 2.8<br>(2.4 - 3.2) |
| 60 | 3.2<br>(3 - 3.4) | 3.2<br>(2.8 - 3.6) | 3.4<br>(3.1 - 3.7) | 3.3<br>(2.9 - 3.7) | 2.9<br>(2.5 - 3.2) | 2.3<br>(2.1 - 2.5) | 2.3<br>(2 - 2.7) |
| 70 | 2.7<br>(2.5 - 2.9) | 2.7<br>(2.4 - 3.1) | 2.9<br>(2.6 - 3.1) | 2.8<br>(2.5 - 3.1) | 2.5<br>(2.2 - 2.7) | 2<br>(1.8 - 2.2) | 2<br>(1.7 - 2.3) |
| 80 | 2.4<br>(2.2 - 2.5) | 2.4<br>(2.1 - 2.7) | 2.5<br>(2.3 - 2.8) | 2.5<br>(2.2 - 2.8) | 2.2<br>(1.9 - 2.4) | 1.7<br>(1.5 - 1.9) | 1.8<br>(1.5 - 2) |
| 90 | 2.1<br>(2 - 2.3) | 2.1<br>(1.9 - 2.4) | 2.2<br>(2 - 2.4) | 2.2<br>(1.9 - 2.4) | 1.9<br>(1.7 - 2.1) | 1.5<br>(1.4 - 1.7) | 1.6<br>(1.3 - 1.8) |
| 100 | 1.9<br>(1.8 - 2) | 1.9<br>(1.7 - 2.2) | 2<br>(1.8 - 2.2) | 2<br>(1.7 - 2.2) | 1.7<br>(1.5 - 1.9) | 1.4<br>(1.2 - 1.5) | 1.4<br>(1.2 - 1.6) |

<sup>a</sup>Expected average proportion of ESBL-*E. coli* out of total *E. coli* in the gut in Switzerland is 19.3%; estimate obtained from the mean proportion found in children in Bangladesh in a previous cohort [24].

The model of the relationships between prevalence and percentage of ESBL-*E. coli* in wastewater assumes fate and transport processes are negligible. Investigating the impacts of environmental variables of precipitation and temperature, which might influence fate and transport, showed weak correlations (Spearman’s ρ ranging from -0.36 to 0.45) with ESBL-*E. coli* prevalence in wastewater. Air temperature showed positive correlations with ESBL-*E. coli* percentage in Chur (ρ = 0.45, p < 0.01) and Altenrhein (ρ = 0.4, p < 0.05), as well as with ESBL-*E. coli* loads in Altenrhein (ρ = 0.41, p < 0.05) and Geneva (ρ = 0.43, p < 0.05). Precipitation did not significantly influence ESBL-*E. coli* or total *E. coli* loads (Supplementary Table S6, Figure S4).

One year of weekly ESBL-*E. coli* data allowed us to explore the impact of monitoring frequency on uncertainty in the estimated annual mean percentage of ESBL-*E. coli* (Figure 4). Our analysis, based on the assumption of a consistent annual mean ESBL-*E. coli* percentage, demonstrates that sampling once a week results in a 95% confidence interval width below ±0.25%. Variations exist among WWTPs, notably with Zurich, Lugano, and Chur displaying higher uncertainty in ESBL-*E. coli* percentage compared to other WWTPs at a given sampling frequency.

**Figure 4:**
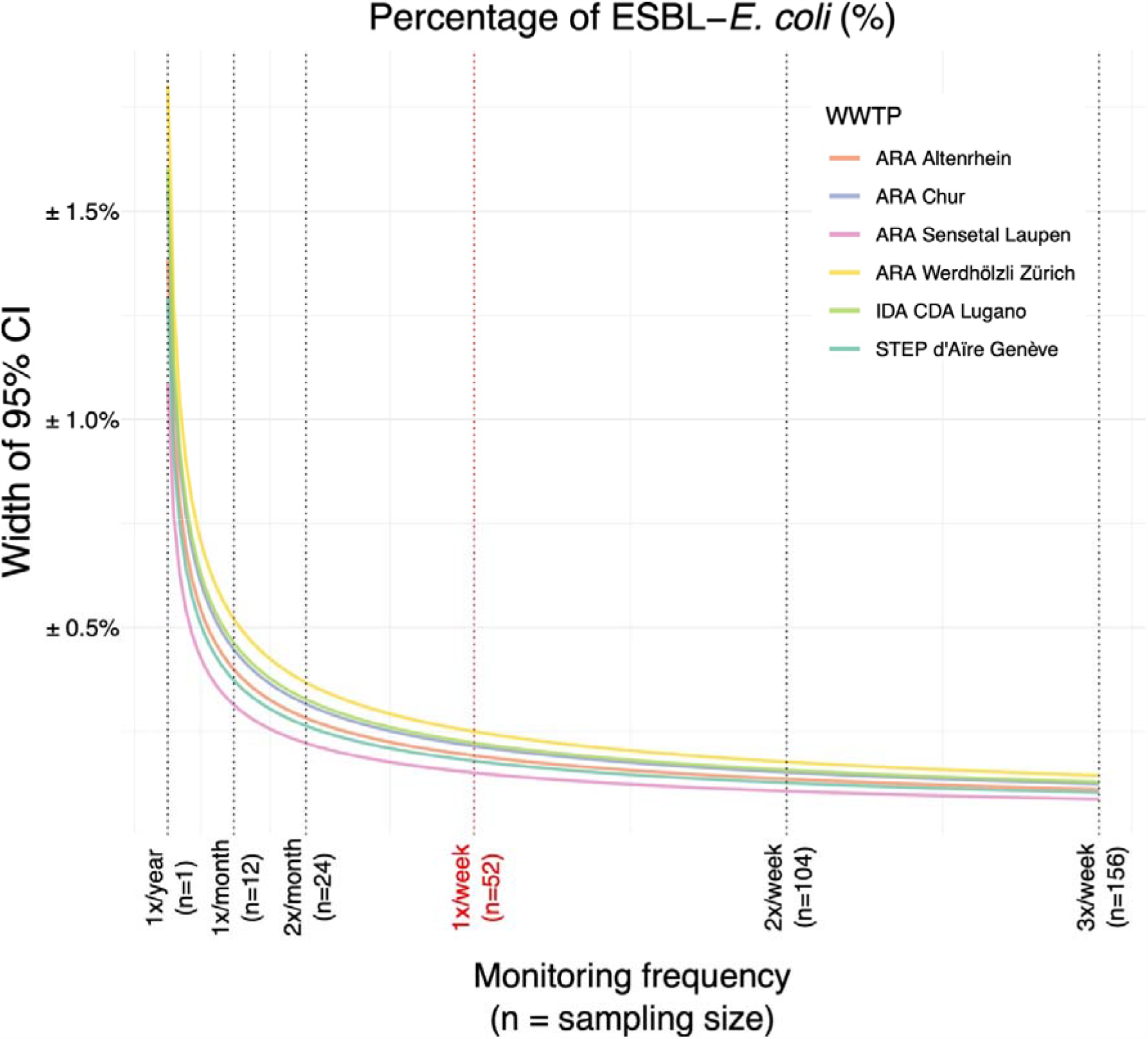
Effect of monitoring frequency on the confidence interval of extended-spectrum β-lactamase-producing *Escherichia coli* percentage in wastewater. CI: confidence interval; *E. coli: Escherichia coli*; ESBL: extended-spectrum β-lactamase; WWTP: wastewater treatment plant The red vertical line highlights the sampling frequency adopted within our study (once a week).

The digital Multiplex Ligation Assay (Figure 5 and Supplementary Table S7) showed that out of 234 ESBL-*E. coli* isolates, 201 (86%) tested positive for at least one target gene family. The positivity rate was consistent across WWTPs, ranging from 72% (n=31 out of 39) in Chur to 92% (n=36 out of 39) in Altenrhein. The most prevalent ESBL-gene family was CTX-M1, identified in 126 isolates (54% of all screened, 95% CI 48%, 59%), present in each WWTP in a minimum of 40% of isolates tested. CTX-M9 and TEM were the second most common, with CTX-M9 detected in 22% (95% CI 14.6%, 29.9%) and TEM also detected in 22% (95% CI 16.9%, 27.5%) of the total ESBL-*E. coli* isolates screened. Less frequent gene families (GES, VIM, PDC) were found in only one isolate each in specific WWTPs (0.4% prevalence, 95% CI 0%, 1.3%). All 18 targeted ESBL-gene families were detected at least once, but there were no instances where all 18 ESBL-gene families were detected within in a single WWTP (Figure 5). Geneva exhibited the highest number of unique gene families (16 out of 18), and Lugano had the least with 10 gene families. Carbapenem resistance, induced by OXA-48, OXA-51, and OXA-231, was infrequently detected in a small number of isolates from all WWTPs except Lugano. No temporal trends in the distribution of ESBL-gene families among isolates were observed (Supplementary Figure S7).

**Figure 5:**
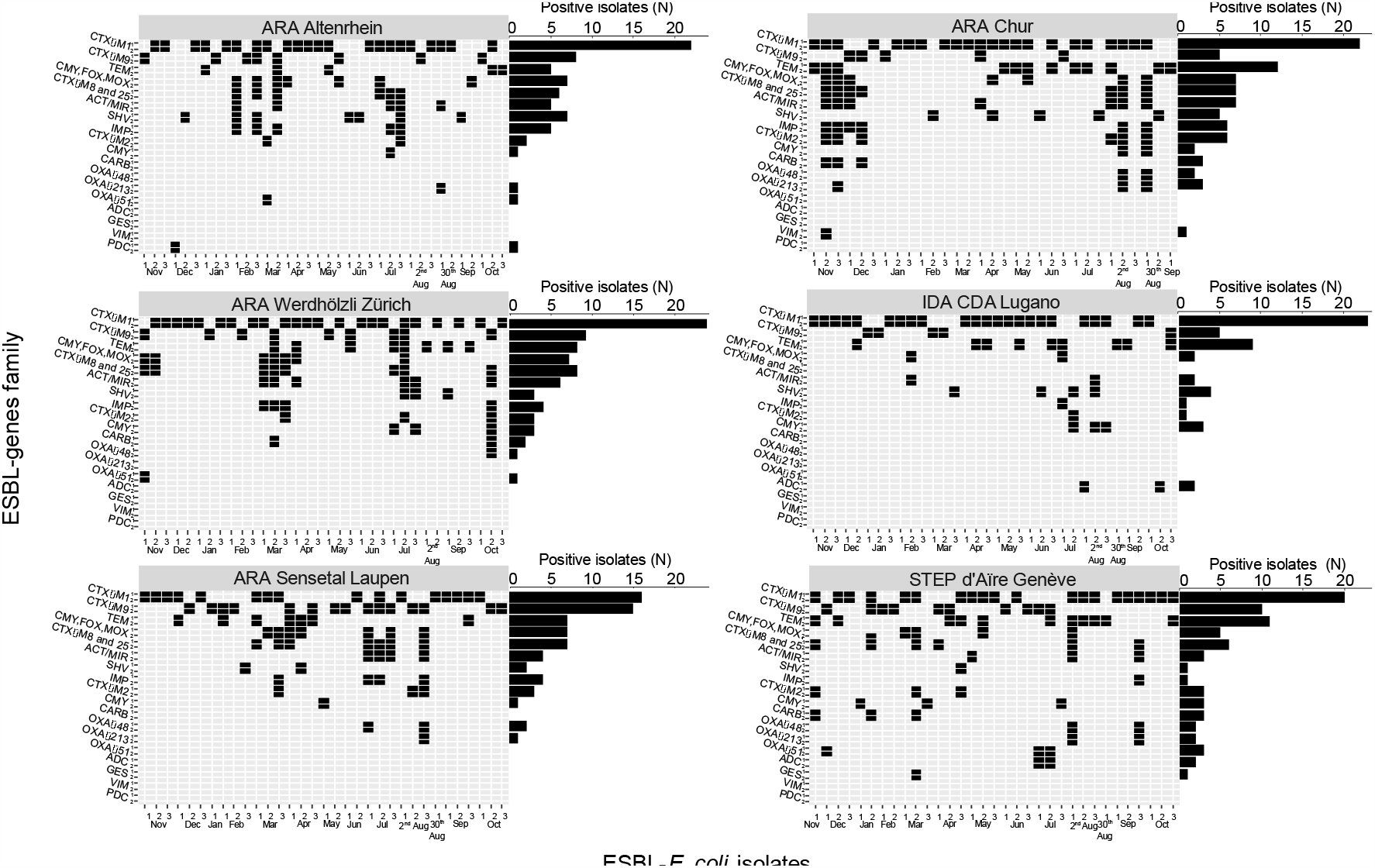
Distribution of 18 extended-spectrum β-lactamase gene families among 201 ESBL-*E. coli* isolates isolated from wastewater, Switzerland, November 2021 – October 2022. *E. coli: Escherichia coli*; ESBL: extended-spectrum β-lactamase; N: number of isolates positive to the gene family Screening of 234 ESBL-*E. coli* isolates (x-axis) for 18 β-lactamase gene families (y-axis) using dMLA with a 36-probe pair mixture (two for each family; named 1 and 2 on the y-axis). Black rectangles indicate presence of the ESBL-gene family in the ESBL-*E. coli* isolate. ESBL-*E. coli* isolates are grouped by wastewater treatment plants (WWTP) and arranged by sampling date (November 2021 to October 2022). Horizontal barplots on the side indicate the number (N) of ESBL-Ec positive to each ESBL-gene family in the respective WWTP. Only strains positive for at least one gene are shown on the heatmap; meaning that both probe-pairs of that gene family were detected in the isolate. 33 strains were negative for all genes and are not represented.

## Discussion

Our results reveal widespread dissemination of ESBL-*E. coli* in Swiss wastewater, with a median concentration of 6.8x10^2^ CFUs/ml and corresponding loads of 2.1x10^8^ CFUs/(person-day). This aligns with previous concentration estimates in Germany (7.6x10^2^ CFUs/ml) [25] and the canton of Basel, Switzerland (9.5x10^1^ CFUs/ml) [13]. Across the six Swiss wastewater treatment plants (WWTPs), we observed a median overall ESBL-*E. coli* percentage of 1.6% (IQR = 0.89), consistent with global rates reported in Sweden (2.3%) [5] and Japan (2.2%) [26]. European data ranged from 1.6% in Greece to 4.4% in Germany, with other countries falling in between, including Denmark, Finland, Norway, Belgium, Italy, and Spain [8].

Studies have demonstrated a correlation between ESBL-*E. coli* prevalence in wastewater and clinical isolates from the same geographic area [5,6], and across geographical locations [8]. Our study highlighted significant variations in ESBL-*E. coli* percentages on a spatial scale, indicating potential differences in carriage rates at each location. The larger WWTPs investigated in terms of number of inhabitants connected to the sewage, namely Zurich, Geneva, and Lugano, showed significantly higher ESBL-*E. coli* percentages throughout the sampling period compared to the smallest WWTPs of Chur and Sensetal-Laupen. Similarly, densely populated regions in Germany also showed increased ESBL-*E. coli* percentage in wastewater [25], suggesting that higher population density facilitates ESBL-*E. coli* transmission.

There are other potential causes for higher percentages of ESBL-*E. coli* than population density alone. Zurich and Geneva are also home of to large international airports, associated with increased international travel and food import, which are known risk factors for the spread of resistant bacterial strains across borders [27,28]. Furthermore, the cities of Zurich and Geneva contain multiple healthcare clinics and hospitals, known to have higher carriage rates compared to community settings [24]. An alternative explanation is that people in larger catchments may have higher proportions of ESBL-*E. coli* in their gut than people in smaller catchments.

Under the assumption that percentage of ESBL-*E. coli* out of total *E. coli* iin the gut is – on average for sufficiently large populations – conserved across Switzerland, wastewater-based estimates may be proxies for carriage rates. Two data points are missing to validate this assumption for Switzerland: the proportion of ESBL-*E. coli* out of total *E. coli* in Swiss carriers and the prevalence of ESBL-*E. coli* carriage in the gut amongst people in Switzerland.When considering the population-averaged median percentage across all WWTPs, and assuming an ESBL-*E. coli* carriage prevalence aligning with the European estimate of 6% [23], the proportion of ESBL-*E. coli* out of total *E. coli* in the gut in the Swiss population would be approximatly 32%. Alternatively, using an average population-based ESBL-*E. coli* prevalence in the gut of 19.3% reported in Bangladesh [24], the ESBL-*E. coli* carriage prevalence in Switzerland ranges between 9.2% and 10.5%, surpassing the European estimate. This discrepancy highlights uncertainty of these estimates in Switzerland. One additional explanation may be the rising global rates of antimicrobial resistance [2]. Our results highlight the potential of wastewater as an indicator of ESBL-*E. coli* carriage within the population, but cohort studies on prevalence rates and shedding loads in Switzerland are necessary to clarify the connection between wastewater and carriage. These studies would also help understanding whether regions with higher ESBL-*E. coli* percentages in wastewater face an elevated risk of ESBL-*E. coli* infections compared to areas with lower percentages. Such research would offer a comprehensive understanding of the relationship between ESBL-*E. coli* presence in wastewater and its potential public health implications.

Observed temporal variations in ESBL-*E. coli* concentrations may be influenced by factors like precipitation and water temperature, which impact runoff and bacterial persistence in sewer transport [29]. We found some association between ESBL-*E. coli* percentage and loads and air temperature, but no clear trend emerged. Previous studies showed similar limited climate influence on wastewater measurements, indicating site-specific variability and sporadic positive correlations between outdoor temperature and antibiotic-resistant bacteria concentration [30,31].

Surveillance of antimicrobial resistance (AMR) using wastewater will require frequent monitoring. The monitoring frequency should represent an optimization of available resources and desired level of estimate of precisions. Our analysis reveals that increasing the sampling frequency to at least once per week results in a 95% CI of approximately ±0.25% for ESBL-*E. coli* percentage. Less frequent sampling, such as monthly monitoring, leads to a 95% CI of about ±0.5%. Sampling frequency is therefore an optimization between budget and logistical constraints (necessitating fewer samples) and acceptable uncertainty in the estimated percentage. Nevertheless, our study highlights that even with reduced sampling, reasonably accurate estimates within a broader confidence interval can still be obtained.

Detection of 18 β-lactamase-encoding genes (ESBL-genes) families in 86% of the 234 ESBL-*E. coli* isolates tested demonstrates high diversity of ESBL-genes in Swiss wastewater. The remaining 14% yielded negative results, potentially due to incomplete gene coverage by the dMLA method [19]. Similar findings occurred in a Polish study, suggesting the presence of other genes in the same molecular class A [32]. Whole-genome sequencing techniques could offer deeper insights into the observed resistance phenotype.

The most prevalent ESBL-gene family in our study was CTX-M-1, found in 54% of screened ESBL-*E. coli* isolates, followed by CTX-M-9 and TEM, present in 22% of isolates. Similar patterns were observed in European studies in France, Netherlands, and Poland (17,32,33). CTX-M-15, a subtype of CTX-M-1, is widely dominant in Europe and commonly associated with the ST131 clone, known for causing antimicrobial-resistant infections in humans globally [34]. This suggests the likely presence of CTX-M enzymes in hospitals or households connected to the investigated wastewater treatment plants.

We found low percentages (2.1% to 3%) of carbapenem-resistant genes, including OXA-48, OXA-51, and OXA-231. Geneva exhibited all three OXA-like gene families, while Lugano had none, indicating geographically constrained circulation. Despite low prevalence, the presence of OXA-like genes in Switzerland raises concerns due to their increasing incidence in clinical settings [35]. VIM and PDC were detected in only 0.4% of screened isolates, aligning with prior observations [36], emphasizing the ability of wastewater monitoring to detect genes with low prevalence and raising questions about their presence in Swiss clinics [37].

In our study, surveyed sites represented large urban areas, covering 14% of Switzerland’s population. Consequently, the obtained ESBL-*E. coli* data may not reflect the entire country, especially in rural or differently populated regions. Additionally, the study detected ESBL-genes only in 86% of isolates, with potential gaps due to protocol limitations. Whole genome sequencing could offer insights for the remaining 14%. Species confirmation for ESBL-*E. coli* was not conducted, but the use of CHROMagar with 99.3% specificity for *E. coli* mitigated biases. Lastly, fluctuations due to bacterial growth or decay within the 48-hour sampling interval might have influenced the results.

In conclusion, the widespread prevalence of ESBL-*E. coli* provides a foundational benchmark useful for informing future trends of AMR population-level carriage in Switzerland. It highlights the potential for investigating the factors driving abundance shifts and their applicability in reducing carriage rates. The estimated nationwide ESBL-*E. coli* carriage prevalence, ranging from 9.2% to 10.5%, surpasses the European estimate, possibly due to escalating global antimicrobial resistance rates. Expanding our research to include other pathogens and cohort studies examining carriage rates would enrich our understanding of AMR dynamics and associated risks for healthcare systems and communities. Furthermore, extending these approaches to communities with decentralized sanitations is needed to gather global insights. Establishing robust connections between our findings and clinical contexts can elucidate the direct impact of AMR gene prevalence on public health. Finally, regular monitoring of antimicrobial resistant pathogens in wastewater across various communities enables the tracking of population-level trends and changes in resistance, and could be used to inform impact of public health interventions.

## Data availability

All data or links to data as well as all scripts are available at our online repository at https://github.com/sheenaconforti/esblec-monitor-ww-21-22.git. Data on precipitation and temperature were obtained from the Federal Office of Meteorology and Climatology MeteoSwiss at https://gate.meteoswiss.ch/idaweb/system/stationList.do (IDAWEB 1.3.5.0 © 2016 MeteoSwiss).

## Acknowledgements

Thanks to Lea Caduff, Charlie Gan, Laura Brülisauer for help processing wastewater samples, Manu Tamminen for help analysing sequencing results, and Tamar Kohn and Christoph Ort for project management and oversight. We extend our acknowledge to Tanja Stadler, her group, and the members of Wastewater-based Surveillance for Infectious diseases and surveillance (WISE) group for their invaluable and insightful inputs for the general discussion. We acknowledge the Natural Sciences and Engineering Research Council of Canada (558161-2021) and the Fonds de Recherche du Québec Nature et technologies (303866) for the postdoctoral fellowship funding of Émile Sylvestre. Finally, thanks to the employees of the wastewater treatment plants IDA CDA Lugano (Ticino), ARA Werdhölzli (Zurich), ARA Chur (Graubünden), ARA Sensetal Laupen (Bern), and STEP d’Aïre Genève (Geneva) for providing samples.

## Authors’ contributions

The author contributions below are according to the CRediT statement. SC: Conceptualization, Methodology, Formal analysis, Investigation, Data Curation, Visualization, Writing-Original Draft, Writing - Review & Editing, Visualization. AH: Methodology, Investigation, Writing - Review & Editing. ES: Methodology, Writing-Review & Editing. TRJ: Conceptualization, Methodology, Resources, Writing – Review & Editing, Supervision, Project administration, Funding acquisition.

