## Supplementary material for "Monitoring ESBL-*Escherichia coli* in Swiss wastewater between November 2021 and November 2022: insights into population carriage": Table S

### Supplementary tables

#### Table S1: Summary of sampling

Total number (N) of collected and missing wastewater samples for each wastewater treatment plant (WWTP) over the 1-year period. The last column indicates the dates of non-sampling from each WWTP. Samples were missed because of cantonal holidays or issues with sampling at the wastewater treatment plant.

| WWTP | Total collected samples (N) | Total missing samples (N) | Dates of missing samples |
|---|---|---|---|
| ARA Altenrhein | 51 | 4 | 2021-12-21; 2021-12-28; 2022-01-04; 2022-01-11 |
| ARA Chur | 51 | 4 | 2021-12-21; 2021-12-28; 2022-01-04; 2022-01-11 |
| STEP d'Aïre Genève | 51 | 4 | 2021-12-21; 2021-12-28; 2022-01-04; 2022-01-11 |
| ARA Sensetal Laupen | 51 | 4 | 2021-12-21; 2021-12-28; 2022-01-04; 2022-01-11 |
| IDA CDA Lugano | 50 | 5 | 2021-12-21; 2021-12-28; 2022-01-04; 2022-01-11; 2022-06-28 |
| ARA Werdhölzli Zürich | 49 | 6 | 2021-12-21; 2021-12-28; 2022-01-04; 2022-01-11; 2022-10-04; 2022-11-29 |

### Table S2: Comparison of ESBL-*E. coli* percentage distributions

Table comparing the lognormal and gamma distributions for ESBL-*E. coli* percentage in the six wastewater treatment plants (WWTPs). The lognormal distribution is described by the parameters μ and σ, while the gamma distribution by the parameters shape (α) and rate (β). The distributions were estimated using a Bayesian model developed by Sylvestre et al., 2020 [21]. D (μ, σ) and D (α, β) indicate the goodness of fit test, while the penalty refers to the complexity of the model. The two distributions were compared using the Deviance Information Criterion (DIC). If the DIC difference is greater than 2, one distribution is significantly better than the other. The last column specifies whether the lognormal or gamma distribution provided a better fit, and if the DIC difference is within ±2, it is labeled as "Same."

| | **Lognormal (μ, $\sigma^2$)** | | | | | **Gamma (α, β)** | | | | | **ΔDIC ((Penalized D (μ, σ)) – (Penalized D (α,β)))** | **Best distribution** |
|---|---|---|---|---|---|---|---|---|---|---|---|---|
| **WWTP** | **μ** | **σ** | **DIC (μ, σ)** | **Penalty** | **Penalized DIC (μ, σ)** | **α** | **β** | **DIC (α,β)** | **Penalty** | **Penalized DIC (α, β)** | | |
| ARA Altenrhein | 0.50 | 0.46 | 90.8 | 2.11 | 93 | 5.56 | 3.1 | 88 | 2.12 | 90.1 | 2.9 | Gamma |
| ARA Chur | 0.25 | 0.51 | 76.5 | 2.15 | 78.7 | 3.97 | 2.73 | 80.1 | 2.17 | 82.3 | -3.6 | Lognormal |
| ARA Werdhölzli Zürich | 0.57 | 0.48 | 89.1 | 2.16 | 91.3 | 4.71 | 2.41 | 90.3 | 2.05 | 92.3 | -1.0 | Same |
| STEP d'Aïre Genève | 0.67 | 0.40 | 95.8 | 2.12 | 97.9 | 7.57 | 3.63 | 89.8 | 2.09 | 91.9 | 6.00 | Gamma |
| IDA CDA Lugano | 0.58 | 0.42 | 88.4 | 2.14 | 90.6 | 6.65 | 3.46 | 84.5 | 2.19 | 86.6 | 4 | Gamma |
| ARA Sensetal Laupen | 0.29 | 0.40 | 62.1 | 2.17 | 64.2 | 6.57 | 4.60 | 62.5 | 2.10 | 64.6 | -0.4 | Same |

**Table S3: Summary statistics of loads, counts and percentage of total and ESBL-*E. coli***

Statistics of *i*) loads of total *E. coli*, *ii*) loads of ESBL-*E. coli*, and *iii*) ESBL-*E. coli* percentage among all samples and within each wastewater treatment plant. The minimum and maximum value, as well as the median (25$^{th}$ to 75$^{th}$ percentile) were calculated for all three variables. $\log_{10}$ mean ± $\log_{10}$ standard deviation (± sd) were calculated for the loads of total *E. coli* and ESBL-*E. coli* only. The gamma arithmetic mean ($\mu_g$) was calculated for the percentage of ESBL-*E. coli* over total *E. coli*. The standard error (SE) of the gamma distributed percentage in each wastewater treatment plant (WWTP) was calculated using the formula $SE = \sqrt{\frac{\alpha}{\beta^2 N}}$. The associated 95% confidence interval (CI) was calculated using the z-distribution.

| | **Total *E. coli*** | | | | | **ESBL-*E. coli*** | | | | | | | |
|---|---|---|---|---|---|---|---|---|---|---|---|---|---|
| | **Loads (CFUs/(person-day))** | | | **Counts (CFUs/mL)** | | **Loads ESBL-*E. coli* (CFUs/(person-day))** | | | **Counts (CFUs/mL)** | | **ESBL-*E. coli* over total *E. coli* (%)** | | |
| | min; max | median (25$^{th}$ to 75$^{th}$) | log10-mean ± sd | min; max | median (25$^{th}$ to 75$^{th}$) | min; max | median (25$^{th}$ to 75$^{th}$) | log10-mean ± sd | min; max | median (25$^{th}$ to 75$^{th}$) | min; max | median (25$^{th}$ to 75$^{th}$) | $\mu_g$ (±95% CI) |
| **All samples** | 4.7x10$^{8}$; 2.5x10$^{11}$ | 1.4x10$^{10}$ (8.4x10$^{9}$-2.2x10$^{10}$) | 10.11 ± 0.38 | 1.5 x 10$^{3}$; 1.9x10$^{5}$ | 4.5x 10$^{4}$ (2.8x10$^{4}$-6.8x10$^{4}$) | 7.5x10$^{6}$; 2.1x10$^{9}$ | 2.1x10$^{8}$ (1.4x10$^{8}$-3.3x10$^{8}$) | 8.30± 0.36 | 4.5x10$^{1}$; 4.2x10$^{3}$ | 6.8x 10$^{2}$ (4.3x10$^{2}$-1.0x10$^{3}$) | 0.36; 5.81 | 1.62 (1.22-2.11) | 1.9 ± 0.1[a] |
| **ARA Altenrhein** | 3.6x10$^{9}$; 9.7x10$^{10}$ | 2.3x10$^{10}$ (1.3x10$^{10}$-2.7x10$^{10}$) | 10.29 ± 0.27 | 8.0 x 10$^{3}$; 1.9x10$^{5}$ | 5.5x 10$^{4}$ (3.3x10$^{4}$-8.5x10$^{4}$) | 7.6x10$^{7}$; 1.5x10$^{9}$ | 3.3x10$^{8}$ (2.0x10$^{8}$-4.3x10$^{8}$) | 8.48± 0.27 | 2.3x10$^{2}$; 4.2x10$^{3}$ | 7.4x 10$^{2}$ (5.2x10$^{2}$-1.2x10$^{3}$) | 0.36; 4.04 | 1.57 (1.27-2.10) | 1.7 ± 0.2 |
| **ARA Chur** | 2x10$^{9}$; 2.5x10$^{11}$ | 1.5x10$^{10}$ (9.9x10$^{9}$-2.1x10$^{10}$) | 10.19 ± 0.38 | 9.5 x 10$^{3}$; 1.1x10$^{5}$ | 5.6x 10$^{4}$ (3.5x10$^{4}$-8.2x10$^{4}$) | 4.9x10$^{7}$; 2.1x10$^{9}$ | 1.8x10$^{8}$ (1.3x10$^{8}$-2.6x10$^{8}$) | 8.28± 0.33 | 1.6x10$^{2}$; 1.5x10$^{3}$ | 5.6x 10$^{2}$ (4.2x10$^{2}$-9.6x10$^{2}$) | 0.39; 5.24 | 1.29 (0.85-1.65) | 1.4 ± 0.2 |
| **STEP d'Aïre Genève** | 1.2x10$^{9}$; 3.9x10$^{10}$ | 1.2x10$^{10}$ (7.2x10$^{9}$-1.9x10$^{10}$) | 10.02 ± 0.38 | 1.3 x 10$^{4}$; 1.4x10$^{5}$ | 4.4x 10$^{4}$ (2.9x10$^{4}$-6.8x10$^{4}$) | 1.8x10$^{7}$; 5.2x10$^{8}$ | 2.7x10$^{8}$ (1.5x10$^{8}$-3.3x10$^{8}$) | 8.30± 0.38 | 2.7x10$^{2}$; 1.9x10$^{3}$ | 8.4x 10$^{2}$ (6.0x10$^{2}$-1.3x10$^{3}$) | 0.43; 3.90 | 2.07 (1.61-2.3) | 2 ± 0.2 |
| **ARA Werdhölzli Zürich** | 7.6x10$^{8}$; 4.2x10$^{10}$ | 1.3x10$^{10}$ (8.2x10$^{9}$-2.2x10$^{10}$) | 10.03 ± 0.44 | 8.5 x 10$^{3}$; 1.5x10$^{5}$ | 3.6x 10$^{4}$ (2.8x10$^{4}$-6.5x10$^{4}$) | 7.5x10$^{6}$; 6.2x10$^{8}$ | 2.7x10$^{8}$ (1.6x10$^{8}$-3.8x1$^{8}$) | 8.28± 0.46 | 1.6x10$^{2}$; 1.8x10$^{3}$ | 7.2x 10$^{2}$ (4.6x10$^{2}$-1.2x10$^{3}$) | 0.49; 5.81 | 1.69 (1.40-2.15) | 1.9 ± 0.3 |
| **IDA CDA Lugano** | 4.7x10$^{8}$; 4x10$^{10}$ | 8.8x10$^{9}$ (8.4x10$^{9}$-1.3x10$^{10}$) | 9.91 ± 0.32 | 1.5 x 10$^{3}$; 8.9x10$^{4}$ | 2.6x 10$^{4}$ (1.5x10$^{4}$-4.2x10$^{4}$) | 1.4x10$^{7}$; 9.6x10$^{8}$ | 1.7x10$^{8}$ (9.1x10$^{7}$-2.5x10$^{8}$) | 8.16± 0.31 | 4.5x10$^{1}$; 1.2x10$^{3}$ | 4.5x 10$^{2}$ (3.1x10$^{2}$-7.1x10$^{2}$) | 0.51; 5.0 | 1.89 (1.46-2.37) | 2 ± 0.2 |

| | | | | | | | | | | | | |
|---|---|---|---|---|---|---|---|---|---|---|---|---|
| **ARA Sensetal Laupen** | $1.3x10^{9}$; $1.5x10^{11}$ | $1.5x10^{10}$ ($1.0x10^{10}$-$2.3x10^{10}$) | 10.19 ± 0.36 | 1.0 x $10^{4}$; $1.0x10^{5}$ | 5.2x $10^{4}$ ($3.4x10^{4}$-$7.0x10^{4}$) | $1.5x10^{7}$; $1.3x10^{9}$ | $2.0x10^{8}$ ($1.4x10^{8}$-$2.5x10^{8}$) | 8.30± 0.31 | $1.5x10^{2}$; $1.7x10^{3}$ | 6.1x $10^{2}$ ($4.5x10^{2}$-$8.2x10^{2}$) | 0.44; 3.13 | 1.26 (0.99-1.66) | 1.4 ± 0.2 |

**[a]** the population-averaged mean proportion of ESBL-*E. coli* detected in Swiss wastewater (1.9 ± 0.1) was derived by multiplying the gamma mean ESBL-*E. coli* percentage in each WWTP by the population connected to that WWTP. The products were then summed across all six WWTPs and divided by the total population (1.23 million individuals), providing a population-averaged measure. The standard error of the weighted-mean ($SEM_W$) was calculated using the following formula by Endlich et *al.,* 1988 [22]:

$SEM_W = \sqrt{\frac{n}{(n-1)(\sum P_i)^2}(\sum P_i X_i - \bar{P}\, X_w)^2 - 2X_w \sum(P_i - \bar{P})(P_i X_i - \bar{P} X_w) + X_w^2 \sum(P_i - \bar{P})^2)}$ , where $n$ is the number of WWTPs, $P_i$ is the population size in each WWTP, $\bar{P}$ is the mean population across WWTPs, $X_i$ is the gamma mean of ESBL-*E. coli* percentage in wastewater in each WWTPs and $X_w$ is the weighted mean. The 95% confidence interval (CI) of the mean was calculated using the z-distribution.

**Table S4: Pairwise comparisons of wastewater treatment plants**

Table showing the results of the pairwise comparisons of *i)* the percentage of ESBL-*E. coli* over total *E. coli*, *ii)* the loads of total *E. coli*, and *iii)* the loads of ESBL-*E. coli* by wastewater treatment plant (WWTP) using the Dunn's test with Bonferroni adjustment. The mean differences (μ) between the WWTPs are shown in the cells (e.g., μ of ESBL-*E. coli* percentage between ARA Altenrhein and ARA Chur is positive, indicating that percentage of ESBL-*E. coli* over total *E. coli* is higher in ARA Altenrhein). The adjusted p-values are considered significant and indicated by * for $p \leq 0.025$.

| Kruskal-Wallis: $p < 0.001$, $X^2 = 48.3$ | | | | | | |
|---|---|---|---|---|---|---|
| **Pairwise comparison** | **ESBL-*E. coli* percentage** | | **Loads total *E. coli*** | | **Loads ESBL-*E. coli*** | |
| | **Mean differences (μ)** | **Significance (adj. *p*-Value)** | **Mean differences (μ)** | **Significance (adj. *p*-Value)** | **Mean differences (μ)** | **Significance (adj. *p*-Value)** |
| ARA Altenrhein – ARA Chur | 2.7 | 0.052 | 2.4 | 0.136 | 4.23 | **0.0002*** |
| ARA Altenrhein – ARA Sensetal Laupen | 2.6 | 0.068 | 2.1 | 0.271 | 3.7 | **0.002*** |
| ARA Altenrhein – ARA Werdhölzli Zürich | -1 | 1 | 3.1 | **0.0158*** | 1.9 | 0.4621 |
| ARA Altenrhein – IDA CDA Lugano | -1.7 | 0.635 | 6.1 | **< 0.0001*** | 5.3 | **< 0.0001*** |
| ARA Altenrhein – STEP d'Aïre Genève | -2.4 | 0.11 | 3.8 | **0.0009*** | 2.1 | 0.284 |
| ARA Chur – ARA Sensetal Laupen | -0.1 | 1 | -0.3 | 1 | -0.6 | 1 |
| ARA Chur – ARA Werdhölzli Zürich | -3.6 | **0.002*** | 0.8 | 1 | -2.3 | 0.1484 |
| ARA Chur – IDA CDA Lugano | -4.4 | **0.0001*** | 3.7 | **0.0016*** | 1.1 | 1 |
| ARA Chur – STEP d'Aïre Genève | -5.2 | **< 0.0001*** | 1.5 | 1 | -2.2 | 0.2282 |
| ARA Sensetal Laupen – ARA Werdhölzli Zürich | -3.6 | **0.0029*** | 1 | 1 | -1.8 | 0.6038 |
| ARA Sensetal Laupen – IDA CDA Lugano | -4.3 | **0.0001*** | 4 | **0.0005*** | 1.7 | 0.7121 |
| ARA Sensetal Laupen – STEP d'Aïre Genève | -5.1 | **< 0.0001*** | 1.7 | 0.6132 | -1.6 | 0.8625 |
| ARA Werdhölzli Zürich – IDA CDA Lugano | -0.7 | 1 | 2.9 | 0.0299 | 3.4 | **0.0052*** |
| ARA Werdhölzli Zürich – STEP d'Aïre Genève | -1.4 | 1 | 0.7 | 1 | 0.2 | 1 |
| IDA CDA Lugano – STEP d'Aïre Genève | -0.7 | 1 | -2.3 | 0.1854 | -3.2 | **0.009*** |

**Table S5: Pairwise comparisons of months within wastewater treatment plants**

Table showing the results of the pairwise comparisons by month in each wastewater treatment plant (WWTP) of *i*) the loads of total *E. coli*, and *ii*) the loads of ESBL-*E. coli*, and *iii*) the percentage of ESBL-*E. coli* over total *E. coli*. Pairwise comparisons were performed with the Dunn's test with Bonferroni adjustment. The significant p-values are indicated by * for p ≤ 0.025.

| **ARA Altenrhein – Loads of total *E. coli* p-values** | | | | | | | | | | | | | |
|---|---|---|---|---|---|---|---|---|---|---|---|---|---|
| | Nov.21 | Dec.21 | Jan.22 | Feb.22 | Mar.22 | Apr.22 | May.22 | Jun.22 | Jul.22 | Aug.22 | Sep.22 | Oct.22 | Nov.22 |
| Nov.21 | | | | | | | | | | | | | |
| Dec.21 | 1.00 | | | | | | | | | | | | |
| Jan.22 | 1.00 | 1.00 | | | | | | | | | | | |
| Feb.22 | 1.00 | 1.00 | 1.00 | | | | | | | | | | |
| Mar.22 | 1.00 | 1.00 | 1.00 | 1.00 | | | | | | | | | |
| Apr.22 | 1.00 | 1.00 | 1.00 | 1.00 | 1.00 | | | | | | | | |
| May.22 | 1.00 | 1.00 | 1.00 | 1.00 | 1.00 | 1.00 | | | | | | | |
| Jun.22 | 1.00 | 1.00 | 1.00 | 1.00 | 1.00 | 1.00 | 1.00 | | | | | | |
| Jul.22 | 1.00 | 1.00 | 1.00 | 1.00 | 1.00 | 1.00 | 1.00 | 1.00 | | | | | |
| Aug.22 | 1.00 | 1.00 | 1.00 | 1.00 | 1.00 | 1.00 | 0.17 | 1.00 | 1.00 | | | | |
| Sep.22 | 0.68 | 0.77 | 1.00 | 1.00 | 1.00 | 0.32 | **0.0028*** | 1.00 | 1.00 | 1.00 | | | |
| Oct.22 | 1.00 | 1.00 | 1.00 | 1.00 | 1.00 | 1.00 | 0.86 | 1.00 | 1.00 | 1.00 | 1.00 | | |
| Nov.22 | 1.00 | 1.00 | 1.00 | 1.00 | 1.00 | 0.83 | **0.0084*** | 1.00 | 1.00 | 1.00 | 1.00 | 1.00 | |

| **ARA Chur – Loads of total *E. coli* p-values** | | | | | | | | | | | | | |
|---|---|---|---|---|---|---|---|---|---|---|---|---|---|
| | Nov.21 | Dec.21 | Jan.22 | Feb.22 | Mar.22 | Apr.22 | May.22 | Jun.22 | Jul.22 | Aug.22 | Sep.22 | Oct.22 | Nov.22 |
| Nov.21 | | | | | | | | | | | | | |
| Dec.21 | 1.00 | | | | | | | | | | | | |
| Jan.22 | 1.00 | 1.00 | | | | | | | | | | | |
| Feb.22 | 1.00 | 1.00 | 1.00 | | | | | | | | | | |
| Mar.22 | 1.00 | 1.00 | 1.00 | 1.00 | | | | | | | | | |
| Apr.22 | 1.00 | 0.36 | 1.00 | 1.00 | 1.00 | | | | | | | | |
| May.22 | 0.27 | 0.09 | 1.00 | 0.57 | 1.00 | 1.00 | | | | | | | |
| Jun.22 | 1.00 | 1.00 | 1.00 | 1.00 | 1.00 | 1.00 | 1.00 | | | | | | |
| Jul.22 | 1.00 | 1.00 | 1.00 | 1.00 | 1.00 | 1.00 | 1.00 | 1.00 | | | | | |
| Aug.22 | 1.00 | 1.00 | 1.00 | 1.00 | 1.00 | 1.00 | 0.71 | 1.00 | 1.00 | | | | |
| Sep.22 | 1.00 | 1.00 | 1.00 | 1.00 | 1.00 | 1.00 | 0.30 | 1.00 | 1.00 | 1.00 | | | |
| Oct.22 | 1.00 | 1.00 | 1.00 | 1.00 | 1.00 | 1.00 | 1.00 | 1.00 | 1.00 | 1.00 | 1.00 | | |
| Nov.22 | 1.00 | 1.00 | 1.00 | 1.00 | 1.00 | 0.30 | 0.04 | 1.00 | 1.00 | 1.00 | 1.00 | 1.00 | |

| **STEP d'Aïre Genève – Loads of total *E. coli* p-values** | | | | | | | | | | | | | |
|---|---|---|---|---|---|---|---|---|---|---|---|---|---|
| | Nov.21 | Dec.21 | Jan.22 | Feb.22 | Mar.22 | Apr.22 | May.22 | Jun.22 | Jul.22 | Aug.22 | Sep.22 | Oct.22 | Nov.22 |
| Nov.21 | | | | | | | | | | | | | |
| Dec.21 | 1.00 | | | | | | | | | | | | |
| Jan.22 | 1.00 | 1.00 | | | | | | | | | | | |
| Feb.22 | 1.00 | 1.00 | 1.00 | | | | | | | | | | |
| Mar.22 | 1.00 | 1.00 | 1.00 | 1.00 | | | | | | | | | |
| Apr.22 | 1.00 | 1.00 | 1.00 | 1.00 | 1.00 | | | | | | | | |
| May.22 | 1.00 | 1.00 | 1.00 | 1.00 | 1.00 | 1.00 | | | | | | | |
| Jun.22 | 1.00 | 1.00 | 1.00 | 1.00 | 1.00 | 1.00 | 1.00 | | | | | | |
| Jul.22 | 1.00 | 1.00 | 1.00 | 1.00 | 1.00 | 1.00 | 1.00 | 1.00 | | | | | |
| Aug.22 | 0.79 | 1.00 | 1.00 | 1.00 | 0.31 | 0.55 | 1.00 | 1.00 | 1.00 | | | | |

| | | | | | | | | | | | | | |
|---|---|---|---|---|---|---|---|---|---|---|---|---|---|
| Sep.22 | 0.04 | 1.00 | 1.00 | 0.24 | **0.0079*** | **0.0201*** | 0.06 | 1.00 | 1.00 | 1.00 | | | |
| Oct.22 | 0.50 | 1.00 | 1.00 | 1.00 | 0.19 | 1.00 | 0.90 | 1.00 | 1.00 | 1.00 | 1.00 | | |
| Nov.22 | 0.03 | 1.00 | 1.00 | 0.17 | **0.0035*** | **0.0108*** | 0.03 | 1.00 | 1.00 | 1.00 | 1.00 | 1.00 | |
| **ARA Sensetal Laupen – Loads of total *E. coli* p-values** | | | | | | | | | | | | | |
| | Nov.21 | Dec.21 | Jan.22 | Feb.22 | Mar.22 | Apr.22 | May.22 | Jun.22 | Jul.22 | Aug.22 | Sep.22 | Oct.22 | Nov.22 |
| Nov.21 | | | | | | | | | | | | | |
| Dec.21 | 1.00 | | | | | | | | | | | | |
| Jan.22 | 1.00 | 1.00 | | | | | | | | | | | |
| Feb.22 | 1.00 | 1.00 | 1.00 | | | | | | | | | | |
| Mar.22 | 1.00 | 1.00 | 1.00 | 1.00 | | | | | | | | | |
| Apr.22 | 1.00 | 1.00 | 1.00 | 1.00 | 1.00 | | | | | | | | |
| May.22 | 1.00 | 1.00 | 1.00 | 1.00 | 0.25 | 1.00 | | | | | | | |
| Jun.22 | 1.00 | 1.00 | 1.00 | 1.00 | 1.00 | 1.00 | 1.00 | | | | | | |
| Jul.22 | 1.00 | 1.00 | 1.00 | 1.00 | 1.00 | 1.00 | 1.00 | 1.00 | | | | | |
| Aug.22 | 1.00 | 1.00 | 1.00 | 1.00 | 1.00 | 1.00 | 0.35 | 1.00 | 1.00 | | | | |
| Sep.22 | 1.00 | 1.00 | 1.00 | 1.00 | 1.00 | 1.00 | 0.39 | 1.00 | 1.00 | 1.00 | | | |
| Oct.22 | 1.00 | 1.00 | 1.00 | 1.00 | 1.00 | 1.00 | 0.78 | 1.00 | 1.00 | 1.00 | 1.00 | | |
| Nov.22 | 0.16 | 1.00 | 1.00 | 1.00 | 1.00 | 0.74 | **0.0205*** | 0.60 | 1.00 | 1.00 | 1.00 | 1.00 | |
| **IDA CDA Lugano – Loads of total *E. coli* p-values** | | | | | | | | | | | | | |
| | Nov.21 | Dec.21 | Jan.22 | Feb.22 | Mar.22 | Apr.22 | May.22 | Jun.22 | Jul.22 | Aug.22 | Sep.22 | Oct.22 | Nov.22 |
| Nov.21 | | | | | | | | | | | | | |
| Dec.21 | 1.00 | | | | | | | | | | | | |
| Jan.22 | 1.00 | 1.00 | | | | | | | | | | | |
| Feb.22 | 1.00 | 1.00 | 1.00 | | | | | | | | | | |
| Mar.22 | 1.00 | 1.00 | 1.00 | 1.00 | | | | | | | | | |
| Apr.22 | 1.00 | 1.00 | 1.00 | 1.00 | 1.00 | | | | | | | | |
| May.22 | 1.00 | 1.00 | 1.00 | 1.00 | 1.00 | 1.00 | | | | | | | |
| Jun.22 | 1.00 | 1.00 | 1.00 | 1.00 | 1.00 | 1.00 | 1.00 | | | | | | |
| Jul.22 | 1.00 | 1.00 | 1.00 | 1.00 | 1.00 | 1.00 | 1.00 | 1.00 | | | | | |
| Aug.22 | 1.00 | 1.00 | 1.00 | 1.00 | 1.00 | 1.00 | 1.00 | 1.00 | 1.00 | | | | |
| Sep.22 | 1.00 | 1.00 | 1.00 | 1.00 | 1.00 | 0.56 | 1.00 | 1.00 | 1.00 | 1.00 | | | |
| Oct.22 | 1.00 | 1.00 | 1.00 | 1.00 | 1.00 | 1.00 | 1.00 | 1.00 | 1.00 | 1.00 | 1.00 | | |
| Nov.22 | 1.00 | 1.00 | 1.00 | 1.00 | 1.00 | 1.00 | 1.00 | 1.00 | 1.00 | 1.00 | 1.00 | 1.00 | |
| **ARA Werdhölzli Zürich – Loads of total *E. coli* p-values** | | | | | | | | | | | | | |
| | Nov.21 | Dec.21 | Jan.22 | Feb.22 | Mar.22 | Apr.22 | May.22 | Jun.22 | Jul.22 | Aug.22 | Sep.22 | Oct.22 | Nov.22 |
| Nov.21 | | | | | | | | | | | | | |
| Dec.21 | 1.00 | | | | | | | | | | | | |
| Jan.22 | 1.00 | 1.00 | | | | | | | | | | | |
| Feb.22 | 1.00 | 1.00 | 1.00 | | | | | | | | | | |
| Mar.22 | 1.00 | 0.37 | 1.00 | 1.00 | | | | | | | | | |
| Apr.22 | 1.00 | 1.00 | 1.00 | 1.00 | 1.00 | | | | | | | | |
| May.22 | 1.00 | 0.80 | 1.00 | 1.00 | 1.00 | 1.00 | | | | | | | |
| Jun.22 | 1.00 | 1.00 | 1.00 | 1.00 | 1.00 | 1.00 | 1.00 | | | | | | |
| Jul.22 | 1.00 | 1.00 | 1.00 | 1.00 | 1.00 | 1.00 | 1.00 | 1.00 | | | | | |
| Aug.22 | 0.79 | 0.48 | 1.00 | 1.00 | 0.04 | 0.08 | 0.14 | 1.00 | 1.00 | | | | |
| Sep.22 | 0.28 | 1.00 | 1.00 | 0.56 | **0.0115*** | **0.0236*** | 0.04 | 0.85 | 1.00 | 1.00 | | | |

| | | | | | | | | | | | | |
|---|---|---|---|---|---|---|---|---|---|---|---|---|
| Oct.22 | 1.00 | 1.00 | 1.00 | 1.00 | 1.00 | 1.00 | 1.00 | 1.00 | 1.00 | 1.00 | 1.00 | |
| Nov.22 | 1.00 | 1.00 | 1.00 | 1.00 | 0.37 | 0.48 | 0.80 | 1.00 | 1.00 | 1.00 | 1.00 | 1.00 |

| **ARA Altenrhein – Loads of ESBL-*E. coli* p-values** | | | | | | | | | | | | | |
|---|---|---|---|---|---|---|---|---|---|---|---|---|---|
| | Nov.21 | Dec.21 | Jan.22 | Feb.22 | Mar.22 | Apr.22 | May.22 | Jun.22 | Jul.22 | Aug.22 | Sep.22 | Oct.22 | Nov.22 |
| Nov.21 | | | | | | | | | | | | | |
| Dec.21 | 1.00 | | | | | | | | | | | | |
| Jan.22 | 1.00 | 1.00 | | | | | | | | | | | |
| Feb.22 | 1.00 | 1.00 | 1.00 | | | | | | | | | | |
| Mar.22 | 1.00 | 1.00 | 1.00 | 1.00 | | | | | | | | | |
| Apr.22 | 1.00 | 1.00 | 1.00 | 1.00 | 1.00 | | | | | | | | |
| May.22 | 1.00 | 1.00 | 1.00 | 1.00 | 1.00 | 1.00 | | | | | | | |
| Jun.22 | 1.00 | 1.00 | 1.00 | 1.00 | 1.00 | 1.00 | 1.00 | | | | | | |
| Jul.22 | 1.00 | 1.00 | 1.00 | 1.00 | 1.00 | 1.00 | 1.00 | 1.00 | | | | | |
| Aug.22 | 0.62 | 1.00 | 1.00 | 0.44 | 1.00 | 1.00 | 0.13 | 1.00 | 1.00 | | | | |
| Sep.22 | 1.00 | 1.00 | 1.00 | 1.00 | 1.00 | 1.00 | 0.83 | 1.00 | 1.00 | 1.00 | | | |
| Oct.22 | 1.00 | 1.00 | 1.00 | 1.00 | 1.00 | 1.00 | 0.68 | 1.00 | 1.00 | 1.00 | 1.00 | | |
| Nov.22 | 1.00 | 1.00 | 1.00 | 1.00 | 1.00 | 1.00 | 1.00 | 1.00 | 1.00 | 1.00 | 1.00 | 1.00 | |
| **ARA Chur – Loads of ESBL-*E. coli* p-values** | | | | | | | | | | | | | |
| | Nov.21 | Dec.21 | Jan.22 | Feb.22 | Mar.22 | Apr.22 | May.22 | Jun.22 | Jul.22 | Aug.22 | Sep.22 | Oct.22 | Nov.22 |
| Nov.21 | | | | | | | | | | | | | |
| Dec.21 | 1.00 | | | | | | | | | | | | |
| Jan.22 | 1.00 | 1.00 | | | | | | | | | | | |
| Feb.22 | 1.00 | 1.00 | 1.00 | | | | | | | | | | |
| Mar.22 | 1.00 | 1.00 | 1.00 | 1.00 | | | | | | | | | |
| Apr.22 | 1.00 | 1.00 | 1.00 | 1.00 | 1.00 | | | | | | | | |
| May.22 | 1.00 | 1.00 | 1.00 | 1.00 | 1.00 | 1.00 | | | | | | | |
| Jun.22 | 1.00 | 1.00 | 1.00 | 1.00 | 1.00 | 1.00 | 1.00 | | | | | | |
| Jul.22 | 1.00 | 1.00 | 1.00 | 1.00 | 1.00 | 1.00 | 1.00 | 1.00 | | | | | |
| Aug.22 | 1.00 | 1.00 | 0.48 | 0.83 | 0.60 | 0.07 | 0.04 | 1.00 | 1.00 | | | | |
| Sep.22 | 1.00 | 1.00 | 1.00 | 1.00 | 1.00 | 0.99 | 0.71 | 1.00 | 1.00 | 1.00 | | | |
| Oct.22 | 1.00 | 1.00 | 1.00 | 1.00 | 1.00 | 1.00 | 0.87 | 1.00 | 1.00 | 1.00 | 1.00 | | |
| Nov.22 | 1.00 | 1.00 | 1.00 | 1.00 | 1.00 | 1.00 | 1.00 | 1.00 | 1.00 | 1.00 | 1.00 | 1.00 | |
| **STEP d'Aïre Genève – Loads of ESBL-*E. coli* p-values** | | | | | | | | | | | | | |
| | Nov.21 | Dec.21 | Jan.22 | Feb.22 | Mar.22 | Apr.22 | May.22 | Jun.22 | Jul.22 | Aug.22 | Sep.22 | Oct.22 | Nov.22 |
| Nov.21 | | | | | | | | | | | | | |
| Dec.21 | 1.00 | | | | | | | | | | | | |
| Jan.22 | 1.00 | 1.00 | | | | | | | | | | | |
| Feb.22 | 1.00 | 1.00 | 1.00 | | | | | | | | | | |
| Mar.22 | 1.00 | 1.00 | 1.00 | 1.00 | | | | | | | | | |
| Apr.22 | 1.00 | 1.00 | 1.00 | 1.00 | 1.00 | | | | | | | | |
| May.22 | 1.00 | 1.00 | 1.00 | 1.00 | 1.00 | 1.00 | | | | | | | |
| Jun.22 | 1.00 | 1.00 | 1.00 | 1.00 | 1.00 | 1.00 | 1.00 | | | | | | |
| Jul.22 | 0.14 | 1.00 | 0.99 | 1.00 | 0.21 | 0.64 | 0.49 | 1.00 | | | | | |
| Aug.22 | 0.09 | 1.00 | 1.00 | 0.70 | 0.12 | 0.43 | 0.31 | 1.00 | 1.00 | | | | |
| Sep.22 | 0.11 | 1.00 | 1.00 | 1.00 | 0.17 | 0.52 | 0.39 | 1.00 | 1.00 | 1.00 | | | |
| Oct.22 | 0.25 | 1.00 | 1.00 | 1.00 | 0.42 | 1.00 | 0.90 | 1.00 | 1.00 | 1.00 | 1.00 | | |
| Nov.22 | **0.019*** | 1.00 | 1.00 | 0.18 | **0.0222*** | 0.10 | 1.00 | 1.00 | 1.00 | 1.00 | 1.00 | 1.00 | |

| | | | | | | | | | | | | | |
|---|---|---|---|---|---|---|---|---|---|---|---|---|---|
| **ARA Sensetal Laupen – Loads of ESBL-*E. coli* p-values** | | | | | | | | | | | | | |
| | Nov.21 | Dec.21 | Jan.22 | Feb.22 | Mar.22 | Apr.22 | May.22 | Jun.22 | Jul.22 | Aug.22 | Sep.22 | Oct.22 | Nov.22 |
| Nov.21 | | | | | | | | | | | | | |
| Dec.21 | 1.00 | | | | | | | | | | | | |
| Jan.22 | 1.00 | 1.00 | | | | | | | | | | | |
| Feb.22 | 1.00 | 1.00 | 1.00 | | | | | | | | | | |
| Mar.22 | 1.00 | 1.00 | 1.00 | 1.00 | | | | | | | | | |
| Apr.22 | 1.00 | 1.00 | 1.00 | 1.00 | 1.00 | | | | | | | | |
| May.22 | 1.00 | 1.00 | 1.00 | 1.00 | 0.39 | 1.00 | | | | | | | |
| Jun.22 | 1.00 | 1.00 | 1.00 | 1.00 | 1.00 | 1.00 | 1.00 | | | | | | |
| Jul.22 | 1.00 | 1.00 | 1.00 | 1.00 | 1.00 | 1.00 | 1.00 | 1.00 | | | | | |
| Aug.22 | 0.65 | 0.62 | 1.00 | 0.06 | 1.00 | 1.00 | 0.08 | 0.63 | 1.00 | | | | |
| Sep.22 | 1.00 | 1.00 | 1.00 | 0.87 | 1.00 | 1.00 | 1.00 | 1.00 | 1.00 | 1.00 | | | |
| Oct.22 | 1.00 | 1.00 | 1.00 | 1.00 | 1.00 | 1.00 | 1.00 | 1.00 | 1.00 | 1.00 | 1.00 | | |
| Nov.22 | 1.00 | 1.00 | 1.00 | 1.00 | 1.00 | 1.00 | 0.18 | 1.00 | 1.00 | 1.00 | 1.00 | 1.00 | |
| **IDA CDA Lugano – Loads of ESBL-*E. coli* p-values** | | | | | | | | | | | | | |
| | Nov.21 | Dec.21 | Jan.22 | Feb.22 | Mar.22 | Apr.22 | May.22 | Jun.22 | Jul.22 | Aug.22 | Sep.22 | Oct.22 | Nov.22 |
| Nov.21 | | | | | | | | | | | | | |
| Dec.21 | 1.00 | | | | | | | | | | | | |
| Jan.22 | 1.00 | 1.00 | | | | | | | | | | | |
| Feb.22 | 1.00 | 1.00 | 1.00 | | | | | | | | | | |
| Mar.22 | 1.00 | 1.00 | 1.00 | 1.00 | | | | | | | | | |
| Apr.22 | 1.00 | 1.00 | 0.94 | 1.00 | 1.00 | | | | | | | | |
| May.22 | 1.00 | 1.00 | 1.00 | 1.00 | 1.00 | 1.00 | | | | | | | |
| Jun.22 | 1.00 | 1.00 | 1.00 | 1.00 | 1.00 | 1.00 | 1.00 | | | | | | |
| Jul.22 | 1.00 | 1.00 | 1.00 | 1.00 | 1.00 | 1.00 | 1.00 | 1.00 | | | | | |
| Aug.22 | 1.00 | 1.00 | 1.00 | 1.00 | 1.00 | 1.00 | 1.00 | 1.00 | 1.00 | | | | |
| Sep.22 | 1.00 | 1.00 | 1.00 | 1.00 | 1.00 | 1.00 | 1.00 | 1.00 | 1.00 | 1.00 | | | |
| Oct.22 | 1.00 | 1.00 | 1.00 | 1.00 | 1.00 | 1.00 | 1.00 | 1.00 | 1.00 | 1.00 | 1.00 | | |
| Nov.22 | 1.00 | 1.00 | 1.00 | 1.00 | 1.00 | 1.00 | 1.00 | 1.00 | 1.00 | 1.00 | 1.00 | 1.00 | |
| **ARA Werdhölzli Zürich – Loads of ESBL-Ec p-values** | | | | | | | | | | | | | |
| | Nov.21 | Dec.21 | Jan.22 | Feb.22 | Mar.22 | Apr.22 | May.22 | Jun.22 | Jul.22 | Aug.22 | Sep.22 | Oct.22 | Nov.22 |
| Nov.21 | | | | | | | | | | | | | |
| Dec.21 | 0.48 | | | | | | | | | | | | |
| Jan.22 | 0.43 | 1.00 | | | | | | | | | | | |
| Feb.22 | 1.00 | 1.00 | 0.97 | | | | | | | | | | |
| Mar.22 | 1.00 | 0.20 | 0.18 | 1.00 | | | | | | | | | |
| Apr.22 | 1.00 | 0.67 | 1.00 | 1.00 | 1.00 | | | | | | | | |
| May.22 | 1.00 | 0.68 | 0.61 | 1.00 | 1.00 | 1.00 | | | | | | | |
| Jun.22 | 1.00 | 1.00 | 1.00 | 1.00 | 1.00 | 1.00 | 1.00 | | | | | | |
| Jul.22 | 1.00 | 1.00 | 1.00 | 1.00 | 1.00 | 1.00 | 1.00 | 1.00 | | | | | |
| Aug.22 | 0.28 | 1.00 | 0.60 | 0.71 | 0.06 | 0.37 | 0.33 | 1.00 | 1.00 | | | | |
| Sep.22 | 0.21 | 1.00 | 1.00 | 0.52 | 0.05 | 0.27 | 0.25 | 1.00 | 1.00 | 1.00 | | | |
| Oct.22 | 0.55 | 1.00 | 1.00 | 1.00 | 0.19 | 0.76 | 0.76 | 1.00 | 1.00 | 1.00 | 1.00 | | |
| Nov.22 | 0.50 | 1.00 | 1.00 | 1.00 | 0.14 | 0.68 | 0.66 | 1.00 | 1.00 | 1.00 | 1.00 | 1.00 | |
| **ARA Altenrhein – Percentage of ESBL-*E. coli* p-values** | | | | | | | | | | | | | |
| | Nov.21 | Dec.21 | Jan.22 | Feb.22 | Mar.22 | Apr.22 | May.22 | Jun.22 | Jul.22 | Aug.22 | Sep.22 | Oct.22 | Nov.22 |
| Nov.21 | | | | | | | | | | | | | |
| Dec.21 | 1.00 | | | | | | | | | | | | |

| | | | | | | | | | | | | |
|---|---|---|---|---|---|---|---|---|---|---|---|---|
| Jan.22 | 1.00 | 1.00 | | | | | | | | | | |
| Feb.22 | 1.00 | 1.00 | 1.00 | | | | | | | | | |
| Mar.22 | 1.00 | 1.00 | 1.00 | 1.00 | | | | | | | | |
| Apr.22 | 1.00 | 1.00 | 1.00 | 0.60 | 1.00 | | | | | | | |
| May.22 | 1.00 | 1.00 | 1.00 | 1.00 | 1.00 | 1.00 | | | | | | |
| Jun.22 | 1.00 | 1.00 | 1.00 | 1.00 | 1.00 | 1.00 | 1.00 | | | | | |
| Jul.22 | 1.00 | 1.00 | 1.00 | 1.00 | 1.00 | 1.00 | 1.00 | 1.00 | | | | |
| Aug.22 | 1.00 | 1.00 | 1.00 | 0.97 | 1.00 | 1.00 | 1.00 | 1.00 | 1.00 | | | |
| Sep.22 | 1.00 | 1.00 | 1.00 | 1.00 | 1.00 | 1.00 | 1.00 | 1.00 | 1.00 | 1.00 | | |
| Oct.22 | 1.00 | 1.00 | 1.00 | 1.00 | 1.00 | 1.00 | 1.00 | 1.00 | 1.00 | 1.00 | 1.00 | |
| Nov.22 | 1.00 | 1.00 | 1.00 | 1.00 | 1.00 | 1.00 | 1.00 | 1.00 | 1.00 | 1.00 | 1.00 | 1.00 |

**ARA Chur – Percentage of ESBL-*E. coli* p-values**

| | Nov.21 | Dec.21 | Jan.22 | Feb.22 | Mar.22 | Apr.22 | May.22 | Jun.22 | Jul.22 | Aug.22 | Sep.22 | Oct.22 | Nov.22 |
|---|---|---|---|---|---|---|---|---|---|---|---|---|---|
| Nov.21 | | | | | | | | | | | | | |
| Dec.21 | 1.00 | | | | | | | | | | | | |
| Jan.22 | 1.00 | 1.00 | | | | | | | | | | | |
| Feb.22 | 1.00 | 1.00 | 1.00 | | | | | | | | | | |
| Mar.22 | 1.00 | 1.00 | 1.00 | 1.00 | | | | | | | | | |
| Apr.22 | 1.00 | 1.00 | 1.00 | 1.00 | 1.00 | | | | | | | | |
| May.22 | 1.00 | 0.47 | 1.00 | 0.06 | 1.00 | 1.00 | | | | | | | |
| Jun.22 | 1.00 | 1.00 | 1.00 | 1.00 | 1.00 | 1.00 | 0.68 | | | | | | |
| Jul.22 | 1.00 | 1.00 | 1.00 | 1.00 | 1.00 | 1.00 | 1.00 | 1.00 | | | | | |
| Aug.22 | 1.00 | 0.47 | 1.00 | 0.06 | 1.00 | 1.00 | 1.00 | 0.68 | 1.00 | | | | |
| Sep.22 | 1.00 | 1.00 | 1.00 | 1.00 | 1.00 | 1.00 | 1.00 | 1.00 | 1.00 | 1.00 | | | |
| Oct.22 | 1.00 | 1.00 | 1.00 | 0.87 | 1.00 | 1.00 | 1.00 | 1.00 | 1.00 | 1.00 | 1.00 | | |
| Nov.22 | 1.00 | 1.00 | 1.00 | 1.00 | 1.00 | 1.00 | 0.24 | 1.00 | 1.00 | 0.24 | 1.00 | 1.00 | |

**STEP d'Aïre Genève – Percentage of ESBL-*E. coli* p-values**

| | Nov.21 | Dec.21 | Jan.22 | Feb.22 | Mar.22 | Apr.22 | May.22 | Jun.22 | Jul.22 | Aug.22 | Sep.22 | Oct.22 | Nov.22 |
|---|---|---|---|---|---|---|---|---|---|---|---|---|---|
| Nov.21 | | | | | | | | | | | | | |
| Dec.21 | 1.00 | | | | | | | | | | | | |
| Jan.22 | 1.00 | 1.00 | | | | | | | | | | | |
| Feb.22 | 1.00 | 1.00 | 1.00 | | | | | | | | | | |
| Mar.22 | 1.00 | 1.00 | 1.00 | 1.00 | | | | | | | | | |
| Apr.22 | 0.72 | 1.00 | 1.00 | 1.00 | 1.00 | | | | | | | | |
| May.22 | 1.00 | 1.00 | 1.00 | 1.00 | 1.00 | 1.00 | | | | | | | |
| Jun.22 | 1.00 | 1.00 | 1.00 | 1.00 | 1.00 | 1.00 | 1.00 | | | | | | |
| Jul.22 | 0.23 | 1.00 | 1.00 | 1.00 | 1.00 | 1.00 | 1.00 | 1.00 | | | | | |
| Aug.22 | 0.72 | 1.00 | 1.00 | 1.00 | 1.00 | 1.00 | 1.00 | 1.00 | 1.00 | | | | |
| Sep.22 | 1.00 | 1.00 | 1.00 | 1.00 | 1.00 | 1.00 | 1.00 | 1.00 | 1.00 | 1.00 | | | |
| Oct.22 | 1.00 | 1.00 | 1.00 | 1.00 | 1.00 | 1.00 | 1.00 | 1.00 | 1.00 | 1.00 | 1.00 | | |
| Nov.22 | 1.00 | 1.00 | 1.00 | 1.00 | 1.00 | 0.57 | 1.00 | 1.00 | 0.15 | 0.53 | 1.00 | 1.00 | |

**ARA Sensetal Laupen – Percentage of ESBL-*E. coli* p-values**

| | Nov.21 | Dec.21 | Jan.22 | Feb.22 | Mar.22 | Apr.22 | May.22 | Jun.22 | Jul.22 | Aug.22 | Sep.22 | Oct.22 | Nov.22 |
|---|---|---|---|---|---|---|---|---|---|---|---|---|---|
| Nov.21 | | | | | | | | | | | | | |
| Dec.21 | 1.00 | | | | | | | | | | | | |
| Jan.22 | 1.00 | 1.00 | | | | | | | | | | | |
| Feb.22 | 1.00 | 1.00 | 1.00 | | | | | | | | | | |
| Mar.22 | 1.00 | 1.00 | 1.00 | 1.00 | | | | | | | | | |
| Apr.22 | 1.00 | 0.59 | 1.00 | 0.52 | 1.00 | | | | | | | | |

| | | | | | | | | | | | | | |
|---|---|---|---|---|---|---|---|---|---|---|---|---|---|
| May.22 | 1.00 | 1.00 | 1.00 | 1.00 | 1.00 | 1.00 | | | | | | | |
| Jun.22 | 1.00 | 1.00 | 1.00 | 1.00 | 1.00 | 1.00 | 1.00 | | | | | | |
| Jul.22 | 1.00 | 1.00 | 1.00 | 1.00 | 1.00 | 1.00 | 1.00 | 1.00 | | | | | |
| Aug.22 | 1.00 | 1.00 | 1.00 | 1.00 | 1.00 | 1.00 | 1.00 | 1.00 | 1.00 | | | | |
| Sep.22 | 1.00 | 1.00 | 1.00 | 1.00 | 1.00 | 1.00 | 1.00 | 1.00 | 1.00 | 1.00 | | | |
| Oct.22 | 1.00 | 1.00 | 1.00 | 1.00 | 1.00 | 1.00 | 1.00 | 1.00 | 1.00 | 1.00 | 1.00 | | |
| Nov.22 | 1.00 | 1.00 | 1.00 | 1.00 | 1.00 | 0.23 | 1.00 | 1.00 | 1.00 | 1.00 | 1.00 | 1.00 | |
| **IDA CDA Lugano – Percentage of ESBL-*E. coli* p-values** | | | | | | | | | | | | | |
| | Nov.21 | Dec.21 | Jan.22 | Feb.22 | Mar.22 | Apr.22 | May.22 | Jun.22 | Jul.22 | Aug.22 | Sep.22 | Oct.22 | Nov.22 |
| Nov.21 | | | | | | | | | | | | | |
| Dec.21 | 1.00 | | | | | | | | | | | | |
| Jan.22 | 1.00 | 1.00 | | | | | | | | | | | |
| Feb.22 | 1.00 | 1.00 | 1.00 | | | | | | | | | | |
| Mar.22 | 1.00 | 1.00 | 1.00 | 1.00 | | | | | | | | | |
| Apr.22 | 1.00 | 1.00 | 1.00 | 1.00 | 1.00 | | | | | | | | |
| May.22 | 1.00 | 1.00 | 1.00 | 1.00 | 1.00 | 1.00 | | | | | | | |
| Jun.22 | 1.00 | 1.00 | 1.00 | 1.00 | 1.00 | 1.00 | 1.00 | | | | | | |
| Jul.22 | 1.00 | 1.00 | 1.00 | 1.00 | 1.00 | 1.00 | 1.00 | 1.00 | | | | | |
| Aug.22 | 1.00 | 1.00 | 1.00 | 1.00 | 1.00 | 1.00 | 1.00 | 1.00 | 1.00 | | | | |
| Sep.22 | 1.00 | 1.00 | 1.00 | 1.00 | 1.00 | 1.00 | 1.00 | 1.00 | 1.00 | 1.00 | | | |
| Oct.22 | 1.00 | 1.00 | 1.00 | 1.00 | 1.00 | 1.00 | 1.00 | 1.00 | 1.00 | 1.00 | 1.00 | | |
| Nov.22 | 1.00 | 1.00 | 1.00 | 1.00 | 1.00 | 1.00 | 1.00 | 1.00 | 1.00 | 1.00 | 1.00 | 1.00 | |
| **ARA Werdhölzli Zürich – Percentage of ESBL-*E. coli* p-values** | | | | | | | | | | | | | |
| | Nov.21 | Dec.21 | Jan.22 | Feb.22 | Mar.22 | Apr.22 | May.22 | Jun.22 | Jul.22 | Aug.22 | Sep.22 | Oct.22 | Nov.22 |
| Nov.21 | | | | | | | | | | | | | |
| Dec.21 | 1.00 | | | | | | | | | | | | |
| Jan.22 | 1.00 | 1.00 | | | | | | | | | | | |
| Feb.22 | 1.00 | 1.00 | 1.00 | | | | | | | | | | |
| Mar.22 | 1.00 | 1.00 | 1.00 | 1.00 | | | | | | | | | |
| Apr.22 | 1.00 | 1.00 | 1.00 | 1.00 | 1.00 | | | | | | | | |
| May.22 | 1.00 | 1.00 | 1.00 | 1.00 | 1.00 | 1.00 | | | | | | | |
| Jun.22 | 1.00 | 1.00 | 1.00 | 1.00 | 1.00 | 1.00 | 1.00 | | | | | | |
| Jul.22 | 1.00 | 1.00 | 1.00 | 1.00 | 1.00 | 1.00 | 1.00 | 1.00 | | | | | |
| Aug.22 | 1.00 | 1.00 | 1.00 | 1.00 | 1.00 | 1.00 | 1.00 | 1.00 | 1.00 | | | | |
| Sep.22 | 1.00 | 1.00 | 1.00 | 1.00 | 1.00 | 1.00 | 1.00 | 1.00 | 1.00 | 1.00 | | | |
| Oct.22 | 1.00 | 1.00 | 1.00 | 1.00 | 1.00 | 1.00 | 1.00 | 1.00 | 1.00 | 1.00 | 1.00 | | |
| Nov.22 | 1.00 | 1.00 | 1.00 | 1.00 | 1.00 | 1.00 | 1.00 | 1.00 | 1.00 | 1.00 | 1.00 | 1.00 | |

**Table S6: Correlation analysis with environmental variables**

Table summarizing the Spearman's correlation coefficient between the environmental variables air temperature and precipitations (24h and 96h sum), and the percentage of ESBL-*E. coli* and loads of total and ESBL-*E. coli*. The p-value is Bonferroni-corrected to account for multiple testing. *indicates significant correlations between *E. coli* measures and environmental variables.

| WWTP | Environmental variable | Percentage of ESBL-*E. coli* (%) | | Loads of total *E. coli* (CFUs/person-day) | | Loads of ESBL-*E. coli* (CFUs/person-day) | |
|---|---|---|---|---|---|---|---|
| | | Correlation coefficient (ρ) | p-value | Correlation coefficient (ρ) | p-value | Correlation coefficient (ρ) | p-value |
| ARA Altenrhein | Air temperature (°C) | **0.40** | **0.028*** | 0.07 | 3.765 | **0.41** | **0.018*** |
| | Precipitations (24h sum) | 0.12 | 2.431 | - 0.06 | 3.958 | - 0.01 | 5.566 |
| | Precipitations (96h sum) | 0.23 | 0.636 | 0.07 | 3.822 | 0.20 | 1.026 |
| ARA Chur | Air temperature (°C) | **0.45** | **0.006*** | - 0.23 | 0.596 | 0.17 | 1.446 |
| | Precipitations (24h sum) | - 0.36 | 0.059 | 0.16 | 1.521 | - 0.12 | 2.416 |
| | Precipitations (96h sum) | - 0.21 | 0.814 | 0.13 | 2.105 | - 0.09 | 3.114 |
| STEP d'Aïre Genève | Air temperature (°C) | 0.24 | 0.582 | 0.27 | 0.310 | **0.43** | **0.011*** |
| | Precipitations (24h sum) | - 0.12 | 2.511 | 0.04 | 0.216 | 0.34 | 0.084 |
| | Precipitations (96h sum) | - 0.20 | 0.917 | 0.22 | 0.782 | 0.20 | 1.002 |
| ARA Sensetal Laupen | Air temperature (°C) | 0.32 | 0.131 | 0.03 | 5.084 | 0.25 | 0.493 |
| | Precipitations (24h sum) | -0.28 | 0.272 | 0.06 | 4.157 | - 0.03 | 5.110 |
| | Precipitations (96h sum) | - 0.03 | 4.995 | - 0.03 | 5.052 | 0.04 | 4.734 |
| IDA CDA Lugano | Air temperature (°C) | - 0.15 | 1.806 | - 0.05 | 4.279 | - 0.15 | 1.715 |
| | Precipitations (24h sum) | 0.07 | 3.716 | - 0.09 | 3.315 | - 0.04 | 4.680 |
| | Precipitations (96h sum) | 0.29 | 0.248 | - 0.27 | 0.343 | - 0.11 | 2.592 |
| ARA Werdhölzli Zürich | Air temperature (°C) | 0.16 | 1.682 | 0.19 | 1.270 | 0.21 | 0.861 |
| | Precipitations (24h sum) | - 0.26 | 0.475 | 0.15 | 1.853 | 0.09 | 3.222 |
| | Precipitations (96h sum) | - 0.11 | 2.894 | 0.07 | 3.968 | 0.05 | 4.520 |

**Table S7: ESBL-gene family distribution in ESBL-*E. coli* isolates**

Table showing the number of ESBL-*E. coli* isolates positive to each ESBL-gene family in the six wastewater treatment plants. The total number of ESBL-*E. coli* positive to each ESBL-gene family among all the ESBL-*E. coli* isolates screened is displayed in the last column, together with the respective percentage.

| ESBL-gene family | ARA Altenrhein | ARA Chur | ARA Sensetal Laupen | ARA Werdhölzli Zürich | IDA CDA Lugano | STEP d'Aïre Genève | Total N | Total % (95% CI) |
|---|---|---|---|---|---|---|---|---|
| CTX-M1 | 22 | 22 | 16 | 23 | 23 | 20 | 126 | 53.9% (48.3%, 59.4%) |
| CTX-M9 | 8 | 5 | 15 | 9 | 5 | 10 | 52 | 22.2% (14.6%, 29.9%) |
| TEM | 5 | 12 | 7 | 8 | 9 | 11 | 52 | 22.2% (16.9%, 27.5%) |
| CMY, FOX, MOX | 7 | 7 | 7 | 7 | 2 | 5 | 35 | 15% (10.8%, 19.1%) |
| CTX-M8 and 25 | 6 | 7 | 7 | 8 | 0 | 6 | 34 | 14.5% (8.6%, 20.4%) |
| ACT/MIR | 5 | 7 | 4 | 6 | 2 | 3 | 27 | 11.5% (7.7%, 15.4%) |
| SHV | 7 | 5 | 2 | 3 | 4 | 1 | 22 | 9.4% (5%, 13.8%) |
| IMP | 5 | 6 | 4 | 4 | 1 | 1 | 21 | 9% (4.7%, 13.2%) |
| CTX-M2 | 2 | 6 | 3 | 3 | 1 | 3 | 18 | 7.7% (4.3%, 11.1%) |
| CMY | 1 | 2 | 1 | 3 | 3 | 3 | 13 | 5.6% (3.5%, 7.6%) |
| CARB | 0 | 3 | 0 | 2 | 0 | 3 | 8 | 3.4% (0.3%, 6.5%) |
| OXA-48 | 0 | 2 | 2 | 1 | 0 | 2 | 7 | 3% (1%, 5%) |
| OXA-213 | 1 | 3 | 1 | 0 | 0 | 2 | 7 | 3% (0.6%, 5.4%) |
| OXA-51 | 1 | 0 | 0 | 1 | 0 | 3 | 5 | 2.1% (0%, 4.5%) |
| ADC | 0 | 0 | 0 | 0 | 2 | 2 | 4 | 1.7% (0%, 3.8%) |
| GES | 0 | 0 | 0 | 0 | 0 | 1 | 1 | 0.4% (0%, 1.3%) |
| VIM | 0 | 1 | 0 | 0 | 0 | 0 | 1 | 0.4% (0%, 1.3%) |
| PDC | 1 | 0 | 0 | 0 | 0 | 0 | 1 | 0.4% (0%, 1.3%) |

### Supplementary figures

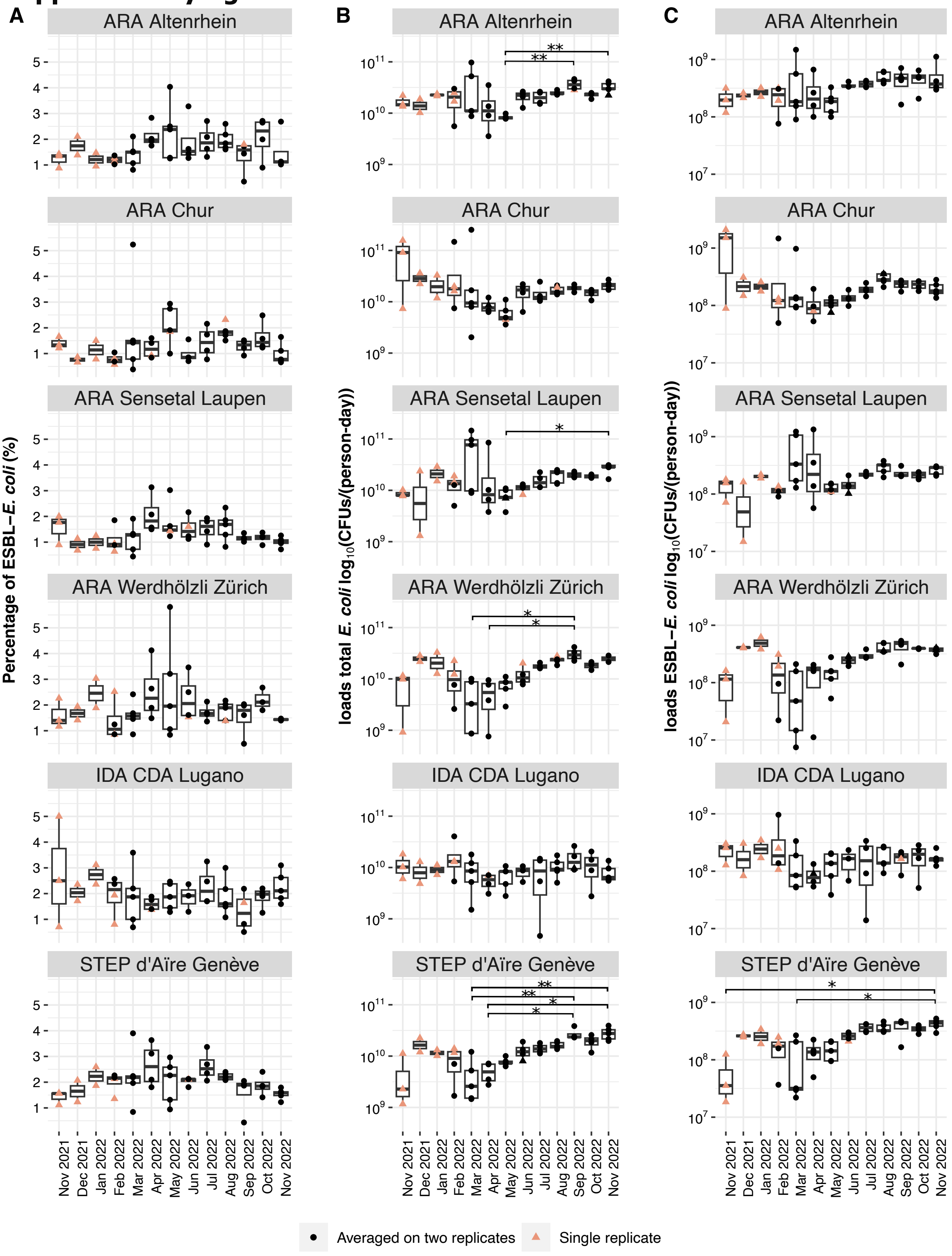

**Figure S1: Monthly distribution of percentage and loads**

Monthly distribution of (A) ESBL-*E. coli* percentage, (B) $\log_{10}$ transformed total *E. coli* loads, and (C) $\log_{10}$ transformed ESBL-*E. coli* loads from November 2021 to November 2022. Black circles represent the averaged value of sample duplicates, and salmon triangles represent the value of the sample single replicate because duplicates were unavailable. * indicates a significant difference with p<0.02 and ** with p<0.001.

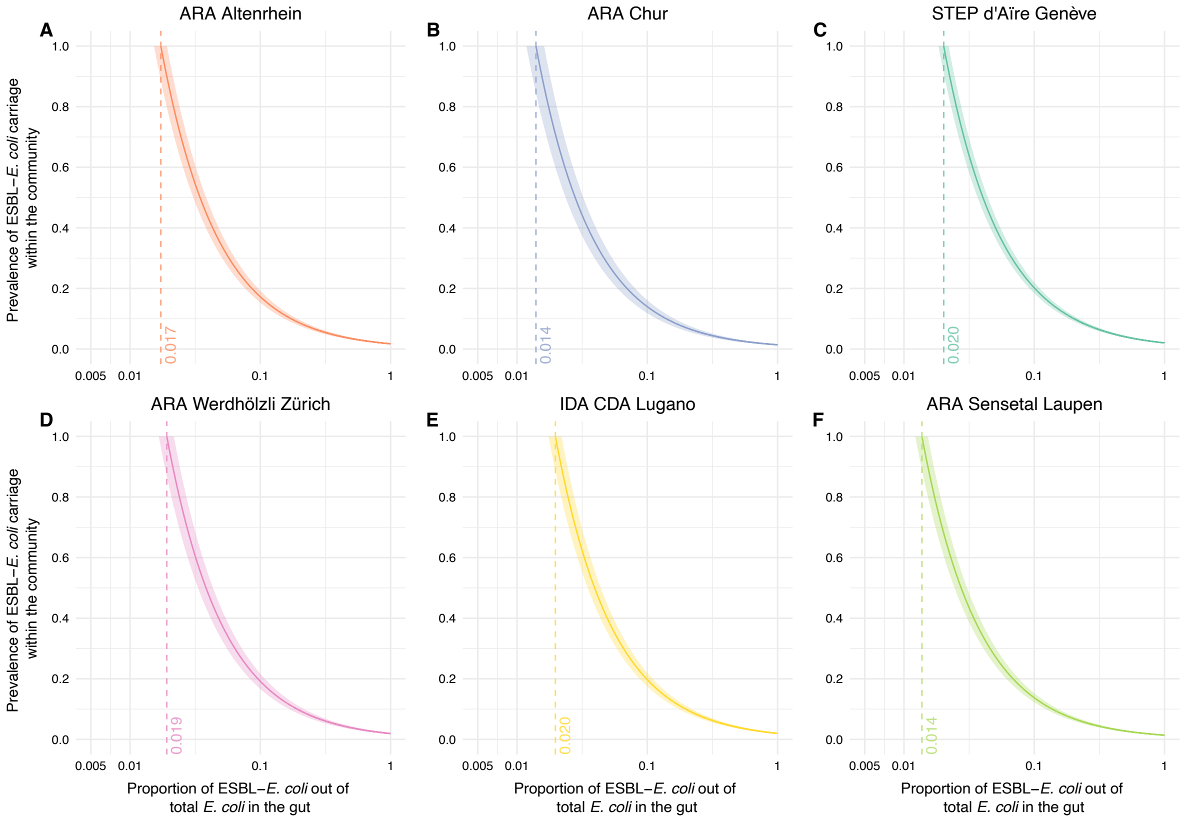

**Figure S2: ESBL-*E. coli* carrier prevalence within wastewater treatment plants**

The figure describes the prevalence of ESBL-*E. coli* carriers ($N_C$) relative to the total population size ($N_T$) as a function of the mean percentage of ESBL-*E. coli* in wastewater ($\overline{\pi}_{WW}$) and the mean proportion of ESBL-*E. coli* relative to the total *E. coli* in the gut discharged by carriers into wastewater ($\overline{\pi}_C$). All panel describe the relationship assuming an overarching estimate of ESBL-*E. coli* out of total *E. coli* in the gut ($\overline{\pi}_C$, x-axis) equal to the ESBL-*E. coli* percentage in wastewater samples ($\overline{\pi}_{WW}$) of the corresponding wastewater treatment plant (WWTP). If the proportion of ESBL-*E. coli* in the gut ($\overline{\pi}_C$) equals the ESBL-*E. coli* percentage in wastewater ($\overline{\pi}_{WW}$), then 100% of people shedding *E. coli* in the catchment of that WWTP carries ESBL-*E. coli* ($\frac{N_C}{N_T}$, y-axis). The shaded areas on the graph represent the 95% confidence interval of the mean percentage ESBL-*E. coli* percentage in wastewater ($\overline{\pi}_{WW}$). The six plots represent the following WWTPs: (A) ARA Altenrhein, (B) ARA Chur, (C) STEP d'Aïre Genève, (D) ARA Werdhölzli Zürich, (E) IDA CDA Lugano, and (F) ARA Sensetal Laupen. Vertical dotted lines indicate the ESBL-*E. coli* percentage detected in wastewater samples ($\overline{\pi}_{WW}$) from the corresponding WWTPs.

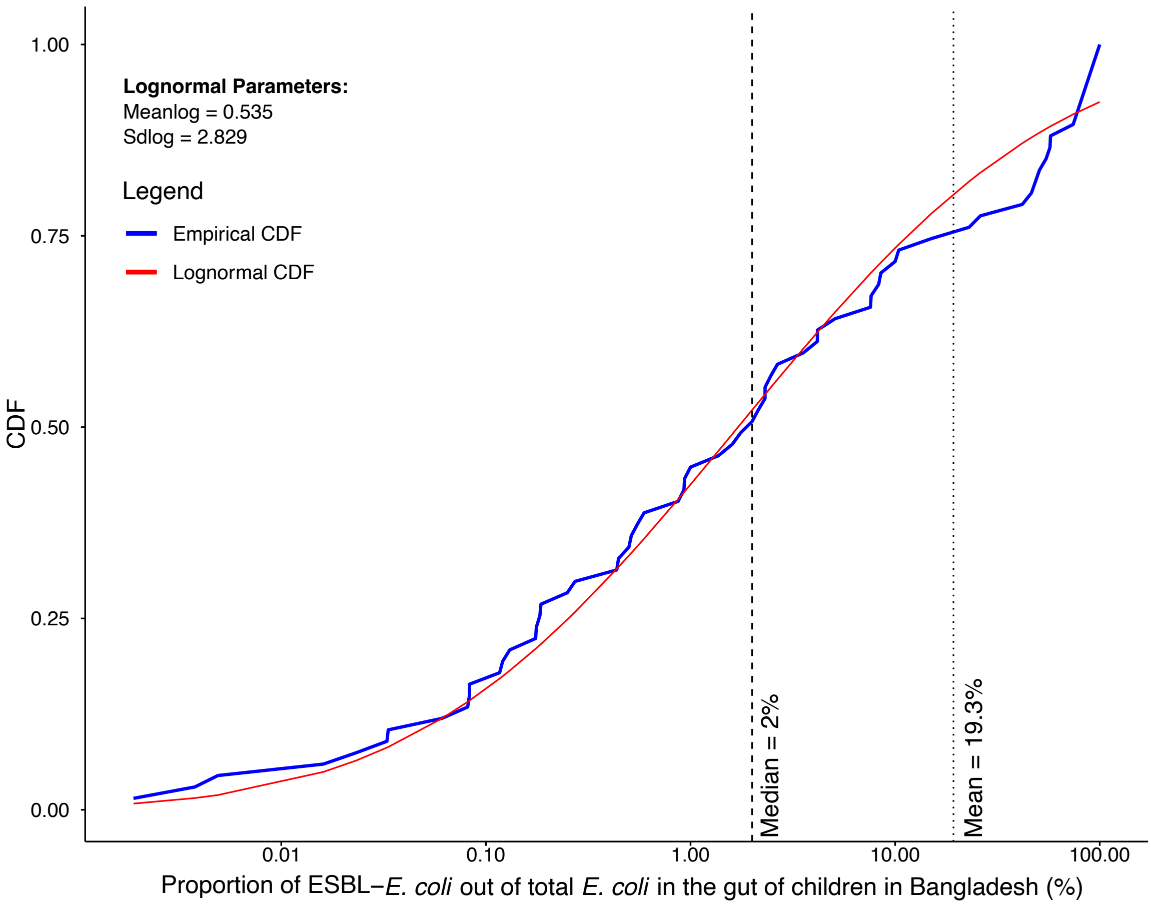

**Figure S3: Distribution of intestinal ESBL-*E. coli* carriage in children of Bangladesh**

This figure displays the distribution of ESBL-*E. coli* proportions relative to total *E. coli* in the gut of 67 children in Bangladesh, sourced from the cohort study of Montealegre et *al.*, 2022 [24]. The values of the logmean and logsd of the lognormal distribution are plotted on the top-left corner of the graph. Notably, the curve shows a mean intestinal carriage of 19.3% and a median of 2%. In our Swiss population study, we utilized the mean to estimate average ESBL-*E. coli* carriers in Switzerland, signifying that the 19.3% prevalence does not imply every individual carrier in Switzerland has an intestinal ESBL-E. coli carriage of 19.3%, but rather reflects the country's average intestinal carriage. The data are available at https://github.com/sheenaconforti/esblec-monitor-ww-21-22.git.

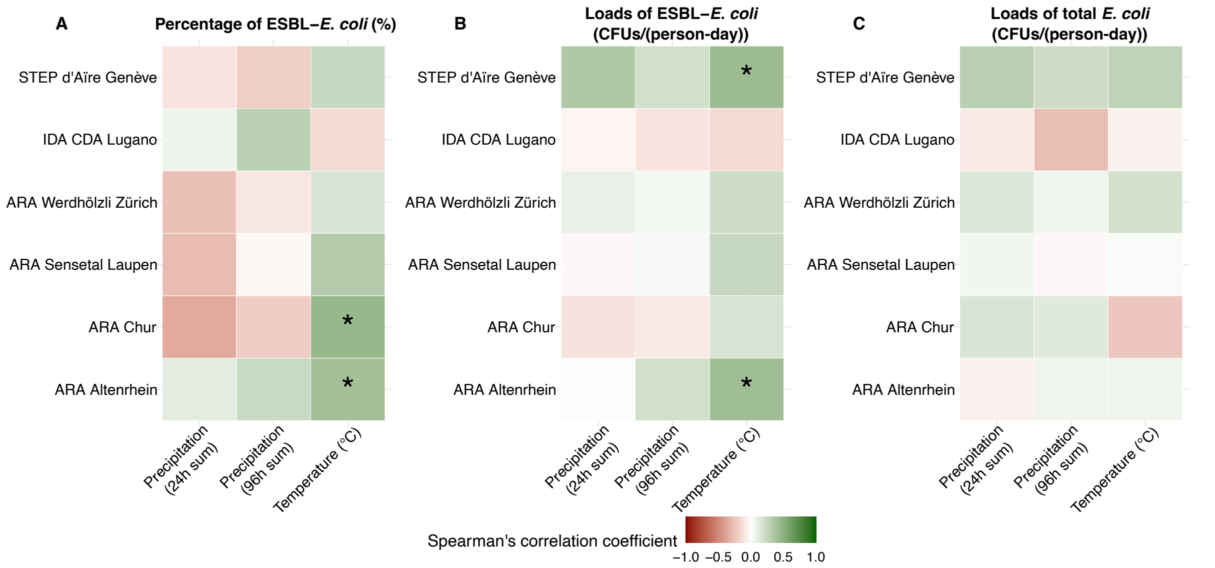

**Figure S4: Correlation analysis with environmental variables**

Heatmap indicating the Spearman's correlation coefficient between the environmental variables (recent precipitation and temperature) and (A) the percentage of ESBL-*E. coli*, (B) the loads of ESBL-*E. coli* and (C) the loads of total *E. coli*. The observations were grouped by wastewater treatment plant (WWTP). The environmental variables considered were *i)* daily total precipitation on the wastewater collection day, *ii)* the cumulative sum of precipitation over 96 hours leading up to and including the wastewater collection, *iii)* the , air temperature data at a height of 2 meters above ground, with a daily average from 6 UTC to 18 UTC. White indicates no correlation; green shades indicate positive correlation and red shades indicate negative correlation. *indicates significant correlation between the variable and the respective WWTP for the observed data. Environmental data were retrieved fromthe Federal Office of Meteorology and Climatology MeteoSwiss at https://gate.meteoswiss.ch/idaweb/system/stationList.do (IDAWEB 1.3.5.0 © 2016 MeteoSwiss). Data for the six meteorological stations located near the investigated WWTPs were downloaded, namely Altenrhein (ARH), Chur (CHU), Mühleberg (MUB), Genève/Cointrin (GVE), Lugano (LUG), and Zürich/Affoltern (REH).

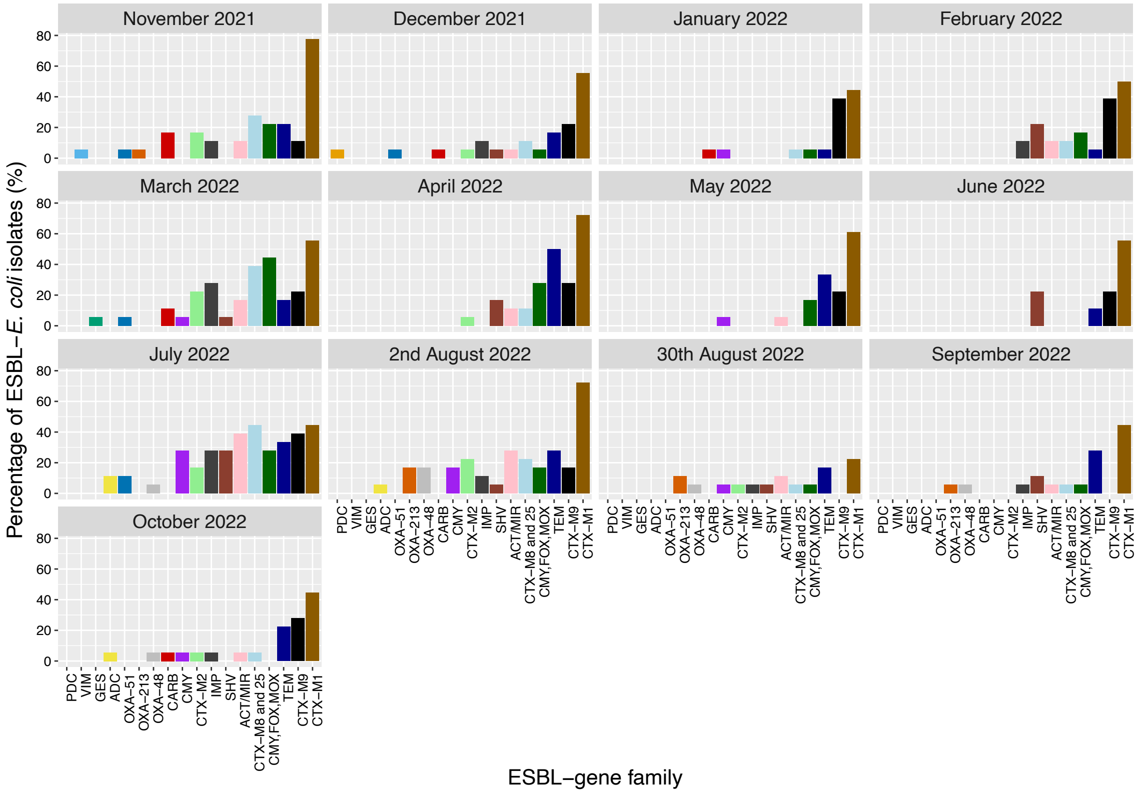

**Figure S5: Temporal distribution of ESBL-gene families in wastewater treatment plants**

Temporal distribution of ESBL-gene families carried by ESBL-*E. coli* isolates among all WWTPs between November 2021 and October 2022.
